# Scalable information extraction from free text electronic health records using large language models

**DOI:** 10.1101/2024.08.08.24311237

**Authors:** Bowen Gu, Vivian Shao, Ziqian Liao, Valentina Carducci, Santiago Romero Brufau, Jie Yang, Rishi J Desai

## Abstract

**Background:** A vast amount of potentially useful information such as description of patient symptoms, family, and social history is recorded as free-text notes in electronic health records (EHRs) but is difficult to reliably extract at scale, limiting their utility in research. This study aims to assess whether an “out of the box” implementation of open-source large language models (LLMs) without any fine-tuning can accurately extract social determinants of health (SDoH) data from free-text clinical notes.

**Methods:** We conducted a cross-sectional study using EHR data from the Mass General Brigham (MGB) system, analyzing free-text notes for SDoH information. We selected a random sample of 200 patients and manually labeled nine SDoH aspects. Eight advanced open-source LLMs were evaluated against a baseline pattern-matching model. Two human reviewers provided the manual labels, achieving 93% inter-annotator agreement. LLM performance was assessed using accuracy metrics for overall, mentioned, and non-mentioned SDoH, and macro F1 scores.

**Results:** LLMs outperformed the baseline pattern-matching approach, particularly for explicitly mentioned SDoH, achieving up to 40% higher Accuracy_mentioned_. openchat_3.5 was the best-performing model, surpassing the baseline in overall accuracy across all nine SDoH aspects. The refined pipeline with prompt engineering reduced hallucinations and improved accuracy.

**Conclusions:** Open-source LLMs are effective and scalable tools for extracting SDoH from unstructured EHRs, surpassing traditional pattern-matching methods. Further refinement and domain-specific training could enhance their utility in clinical research and predictive analytics, improving healthcare outcomes and addressing health disparities.

## BACKGROUND

A vast amount of potentially useful information such as description of patient symptoms, family and social history, is not recorded as structured fields but found in free-text notes from electronic health records (EHRs). Substantial feature engineering is needed to directly extract and analyze the relevant information contained within free-text notes, which is time and resource intensive.^[1–3]^ Therefore, free-text notes have historically been underutilized for research activities.[4]

Various Natural Language Processing (NLP) models have seen developed to extract clinical concepts from EHR notes automatically.[5–6] Traditional rule-based NLP approaches are often constrained by their lack of generalizability across different datasets and settings.[7–8] Machine learning-based NLP models rely heavily on the costly step of manual data annotation for model training, which makes them harder to scale across diverse concepts, particularly in clinical environments where data annotation requires specialized knowledge, cost and confidentiality considerations.[9–11] In contrast, Large Language Models (LLMs), advanced deep learning models that are pre-trained on large volumes of text, provide a scalable alternative for the task of clinical information extraction.[12–14] With their capabilities of zero-shot and few-shot learning, LLMs can extract target clinical information from EHR notes without the need for complex rules creation or extensive data annotation.[15–17]

Social determinants of health (SDoH) of patients are the economic and social conditions that influence individual and group differences in health status.[18–20] They are important for assessing and addressing health disparities, and are critical for clinical interventions and research.[21–23] For instance, quantifying the role SDoH in predisposing patients to adverse health outcomes is often of interest to facilitate development of educational and counseling interventions.[24–25] In epidemiologic investigations, various domains of SDoH are also frequently considered important confounding variables that require adjustment.[26–28] While structured EHRs typically lack explicit recording of SDoH information, it is frequently available in free-text form in notes.[29–31] As variables of universal interest, we selected SDoH as a representative use case where scalable extraction methods like LLMs can be of high value. In the current study, we aimed to assess an “out of the box” implementation of 8 advanced open-source LLMs without any fine-tuning for their capabilities on extracting 9 SDoH from free-text EHRs, and compare their performance with the traditional approach of using basic pattern matching.

## METHODS

### Data Source

We used EHR data from the Mass General Brigham (MGB) system, which is the largest healthcare system in the state of Massachusetts. The sampling frame included a total of 1.2 million individuals with deterministically linked EHRs from MGB to insurance claims data from Medicare and Medicaid for the period of 2007-2020. For this study, we used free text associated with the patients’ social history documentation from progress notes using regular expression matching. Since the patients’ social documentation is added incrementally over time, we only used the most recent social documentation for each patient.

### SDoH Questions and Manual Labels

To identify the SDoH that are frequently reported in social documentation, we first performed a manual review of a random sample of 200 patients’ social documentation and summarized 9 aspects of the patient’s SDoH that appeared in > 5% of the notes. This list included marital status, number of children, employment status, educational status, lifestyle factors (use of tobacco, alcohol, illicit drugs, exercise), and cohabitation status. Of the random sample of the 200 reviewed patients, we split the first 100 patients’ social documentation as the validation set to inform prompt engineering (described below) and the remaining 100 patients’ social documentation as the test set to evaluate performance. Following the classic evaluation framework of LLM evaluation, we converted the SDoH extraction as a question-answer problem. That is, for each of the 9 SDoH characteristics, given the EHR notes, we designed a question and candidate options along with the note text as the LLM input, and let the LLM to select the option from the candidates. The SDoH questions, together with their distribution of candidate options in the validation and the test set, are shown in **Supplementary Table S1**.

For each of the 200 patients, two human reviewers (B.G and V.S) manually labeled the 9 SDoH aspects to one of the quantified choices according to the labeling criteria documentation (**Supplementary**: **Annotation Guide** section). Each reviewer labeled the 200 patients independently. The inter-annotator agreement was calculated based on the total number of the 1800 questions (9 SdoH questions * 200 patients) that the two annotators agreed before discussion among all the 1800 questions, which was 93%. For the inconsistently annotated cases, the two annotators discussed them in detail and reached a consensus. New criteria were also added to the labeling criteria documentation that addressed the causes of these inconsistencies.

### Experiment Settings

We selected 8 well performing open source LLMs on the LLM leaderboard hosted by Hugging Face.[32–41] All LLMs used in this study are publicly available. The details of the LLMs and the links to the models can be found in **Supplementary Table S2**. A copy of the model weights went through the AWQ quantization process to generate the quantized model weights.[42–43] Quantization is a technique of reducing the model size for faster inference in resource limited settings.

### Rule-based Baseline Model

To evaluate the comparative performance of LLMs against a common baseline, we designed a model that used pattern matching to extract the answers to the SDoH questions from the patients’ social documentation. The matching patterns were designed according to the labeling criteria. If a match was found in the patients’ social documentation, the output answer was guaranteed to be one of the choices of the SDoH question. If no match was found in the patients’ social documentation, the output answer was “Not mentioned”. To avoid mismatching (e.g. answer “No” was matched to “Not mentioned” since “No” was a substring of “Not mentioned”), we sorted the choices by their character lengths in descending order and matched the response from the longest choice to the shortest choice and stopped matching if the longer choice was matched. The specific patterns for each SDoH question are shown in **Supplementary Table S3.**

### Pipeline Workflow and prompt engineering

We built two pipelines: a default pipeline and a refined pipeline. The default pipeline was designed to run the LLM to extract the SDoHs from the unstructured social documentation using the default prompt for all SDoH questions. Alternatively, the refined pipeline used different engineered prompts on 3 (Q2, Q6, and Q7) of the 9 SDoH questions that most LLMs struggled with in the validation set experiments. We ran both pipelines to compare the effectiveness of the refinement. An illustration of the two pipelines is shown in **Figure 1**. The “LLM Response Postprocessing” step included implementing a systematic code that uses pattern matching to map the model response to one of the choices of the SDoH questions. The “Auto-Grader” took the refined response from the “LLM Response Postprocessing” procedure and compared it against manual labels. When mapped model response matched exactly with the human label, then the auto-grader considered it as an accurate extraction. Otherwise, the auto-grader considered the LLM extraction inaccurate. The “Model Comparator” step combined the graded model responses from the “Auto-Grader” and provided the grading results in a single chart to formulate the final benchmark.

**Figure 1.**
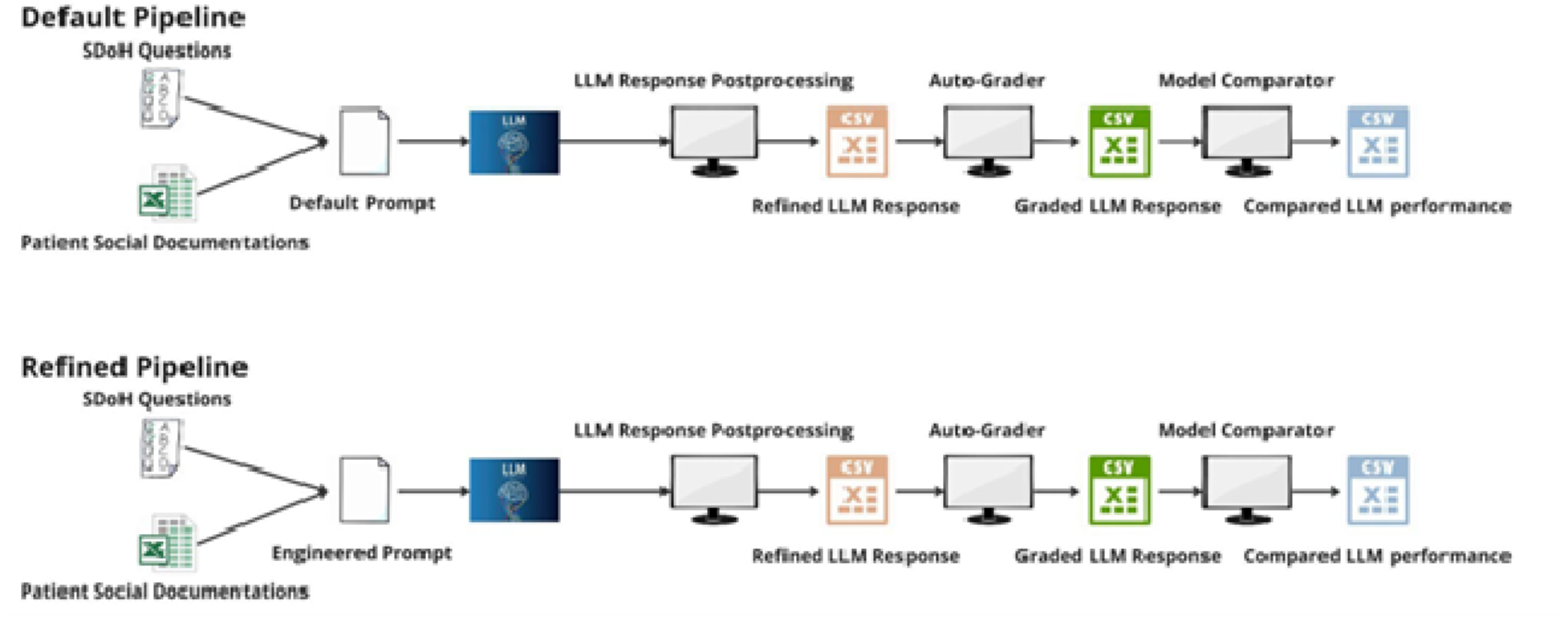
The default and the refined pipeline.

Additional context for the “Default Prompt” and the “Engineered Prompt” is shown in **Supplementary Table S4**, which summarizes 4 types of prompts: default prompts (not including the default secondary prompts), premise prompts, special prompts, and secondary prompts. Th default pipeline only used the default prompts while the refined pipeline used all 4 types of prompts.

### LLM performance evaluation

To evaluate the model performance, we used three metrics in the test set: Accuracy_overall_, Accuracy_mentioned_, and Accuracy_non-mentioned_, which corresponds to the overall accuracy for extractions, the accuracy when a note contained mention of the specific SDoH, and the accuracy when a note did not contain mention of the specific SDoH, respectively. The three accuracies are defined as follows, where the Accuracy_overall_ is a weighted average of the Accuracy_mentioned_ and the Accuracy_non-mentioned_, with the weights dependent on the missingness of the SDoH aspects in the text:

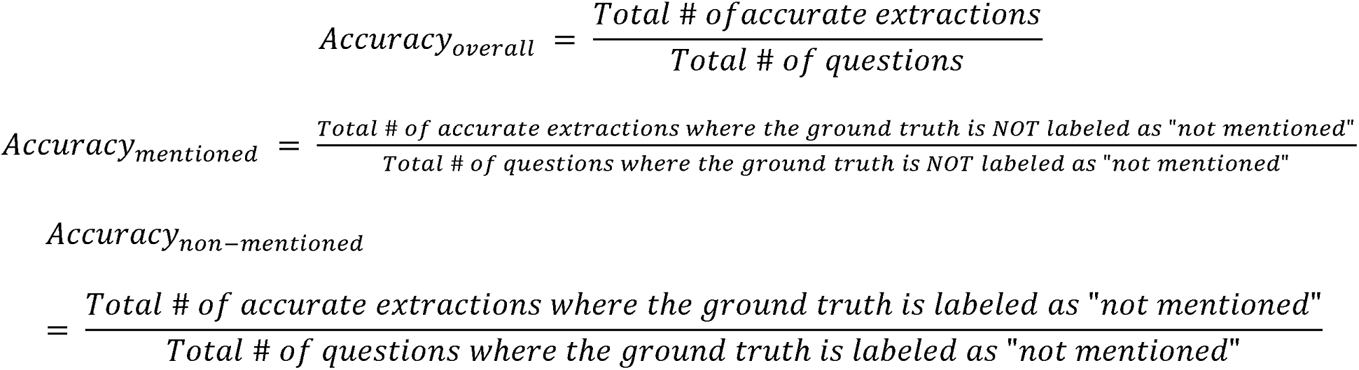

We calculated all three metrics on all 9 SDoH questions. To calculate the confidence interval for these accuracies, we used the Jackknife resampling technique to generate samples for each model accuracy on every question.[44] We then calculated the 95% CI of each accuracy using the samples generated. We assumed t-distribution since our sample size was small (100 per model, per question, per accuracy). For questions that did not meet the premise (e.g. The human label is “not mentioned” for this patient on this SDoH question when trying to calculate Accuracy_mentioned_), we marked them as not applicable for calculations. To evaluate the performance difference between the LLM and the baseline, we also used the Jackknife resampling to generate samples for each model on each question on each accuracy. After that, we performed a two-sided Welch’s t-test on the accuracy differences between the LLM and the baseline using the samples generated from the LLM and the baseline, assuming t-distribution.

Additionally, when the post-processing procedure could not map the LLM response to one of the predefined choices, we defined the response to be invalid and reported the proportion of invalid responses for all models. We further reported the F1 score, calculated as a harmonic mean of precision and recall, using the macro averaging method considering all the SDoH questions as multi-class classification problems. When calculating F1 scores for LLM responses, we combined invalid responses with the ‘not mentioned’ category for each question as a default since invalid responses from LLMs lacked a corresponding manual label.

## RESULTS

### LLM Accuracy

To compare the performance between the LLMs and the baseline, we averaged the model Accuracy_overall_ across the 9 SDoH questions for each LLM and the baseline and did this for the three accuracies defined above (**Figure 2 (a)** and **(b)**). The baseline model achieved a 77.33% average Accuracy_overall_, which was mainly attributable to its high average Accuracy_non-mentioned_. Among all the LLMs, openchat_3.5 was the only LLM that had a significantly better average Accuracy_overall_ than the baseline, it was also the only LLM that outperformed the baseline on each of the 9 SDoH questions on the average Accuracy_overall_. On the other hand, the two Llama-2 models had the worst performance on the average Accuracy_overall_ and were significantly lower than the baseline. The remaining LLMs had comparable average Accuracy_overall_ than the baseline, with the zephyr-7b models performing slightly worse. The baseline, which only had limited patterns to match, had an average Accuracy_mentioned_ of 39.02%; while the majority of the LLMs achieved a higher average Accuracy_mentioned_. Openchat_3.5 achieved over 40% higher of the average Accuracy_mentioned_ than the baseline, which shows its superior capability in extracting information when it is contained in free text. On the other hand, Llama-2-13b-chat was the only LLM that had a significantly worse average Accuracy_mentioned_ compared to the baseline.

**Figure 2(a).**
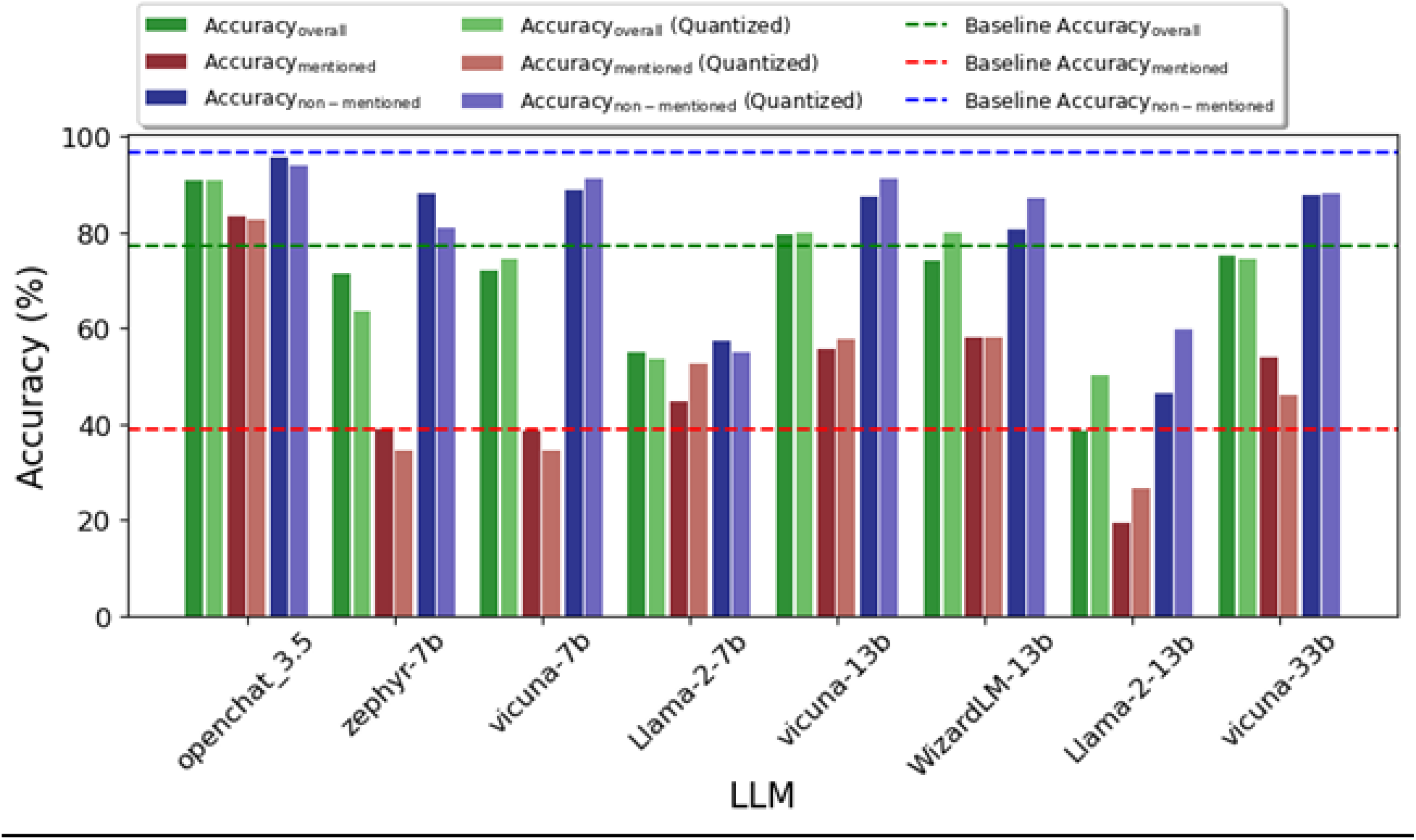
Average accuracies comparison of the LLMs over the baseline (Refined pipeline)

**Figure 2(b).**
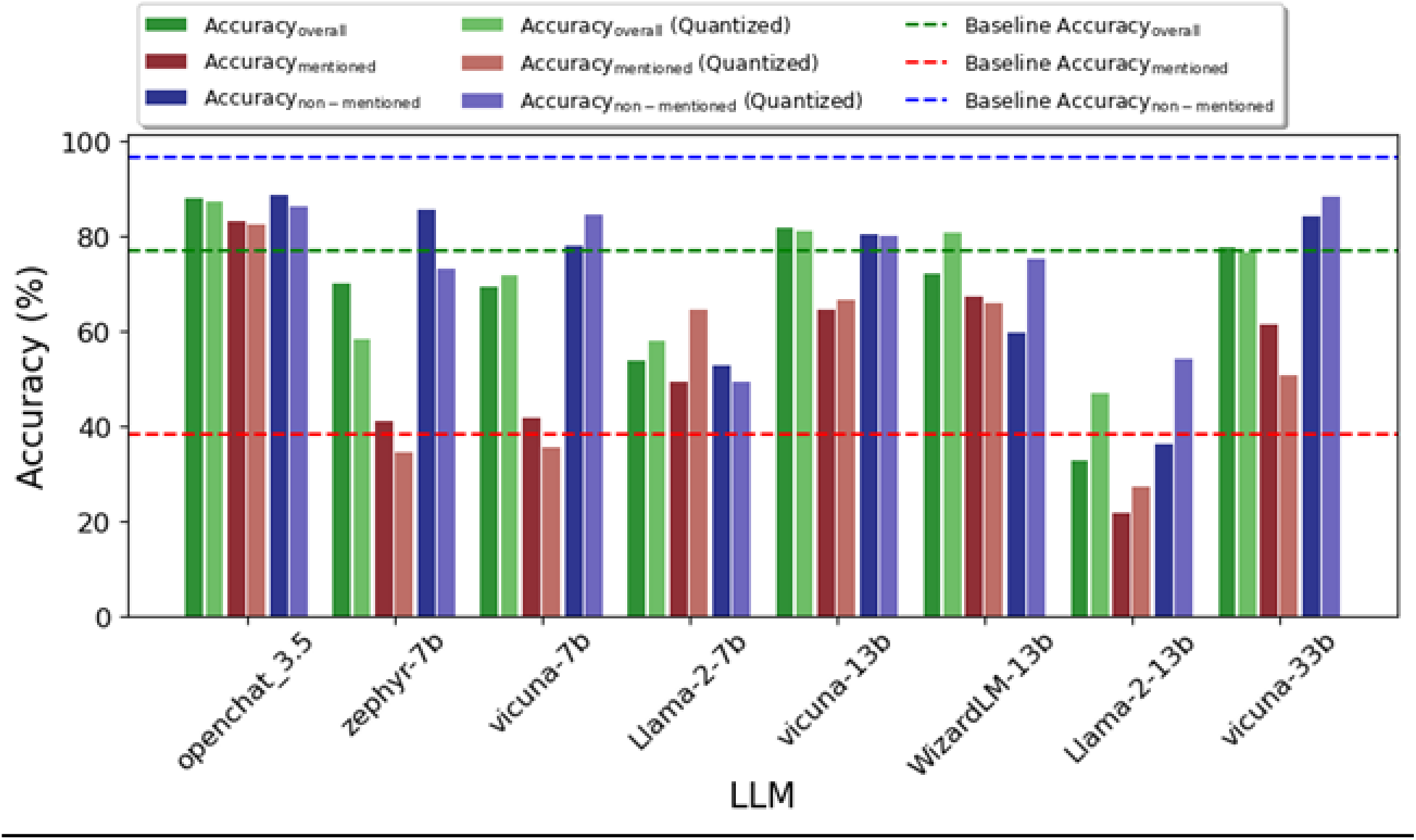
Average accuracies comparison of the LLMs over the baseline (Default pipeline)

The Accuracy_non-mentioned_ is the accuracy that indicates the model’s capability of not producing false information (i.e avoiding “hallucination”).[45–47] The baseline, which reported its responses using pattern matching, had a very slim probability of producing false responses unless the information recorded in the text was internally contradicting. On the other hand, the LLMs had a higher probability of producing false information. We found that the baseline achieved an average Accuracy_non-mentioned_ of 96.66%. This was reasonable as the default response from the baseline is “Not mentioned”. The openchat_3.5 model was the best model in terms of minimizing hallucination. On the contrary, the two models that had poor average Accuracy_non-mentioned_ were the two Llama-2 models.

Compared to the default pipeline, the refined pipeline helped reduce LLM hallucination, but in some instances, this came at a cost of reduced sensitivity to the SDoH features mentioned in the text. This is expected as we used the premise prompt in the refined pipeline, which aims at reducing model hallucinations, but this prompt can make the model more conservative in its responses.

We observed that among all the 9 SDoH questions, Q2 (How many children does the patient have?) and Q7 (What is the patient’s employment status?) were the two questions that most LLMs (except the two Llama 2 models) had a better Accuracy_overall_ than the baseline on both the default and the refined pipeline (**Supplementary Table S6-S7**). This can be explained by **Supplementary Table S8-S9**, which shows that the Accuracy_mentioned_ for the baseline on Q2 and Q7 were very low compared to the LLMs. Besides Q2, and Q7, **Supplementary Table S8-S9** indicates that the baseline model also had very poor Accuracy_mentioned_ on SDoH questions Q3 (Does the patient currently use tobacco?) and Q5 (Does the patient currently use illicit drugs?). This is likely attributable to the fact that the unstructured social history documentation has many ways of expressing the answers on these questions, a simpler rule-based approach misses most of them. **Supplementary Table S10-S11** indicates that the baseline had a much lower probability to produce false positive results compared to LLMs among all 9 SDoH questions. This eliminated the gains in accuracy by LLMs on the Accuracy_mentioned_ and makes the advantage of most LLMs (except the openchat_3.5 model) slim or even negative on the Accuracy_overall_.

### Macro F1 Score

We averaged the model macro F1 scores across the 9 SDoH questions for each LLM and the baseline (**Figure 3).** The baseline had 0.53 average macro F1 for the default pipeline and 0.54 average macro F1 for the refined pipeline. Among all the models, openchat_3.5 was the only LLM that had a clear advantage over the baseline. The two Vicuna models and the WizardLM model had comparable performance as the baseline. The zephyr model and the two Llama 2 models had worse average macro F1 scores than the baseline.

**Figure 3.**
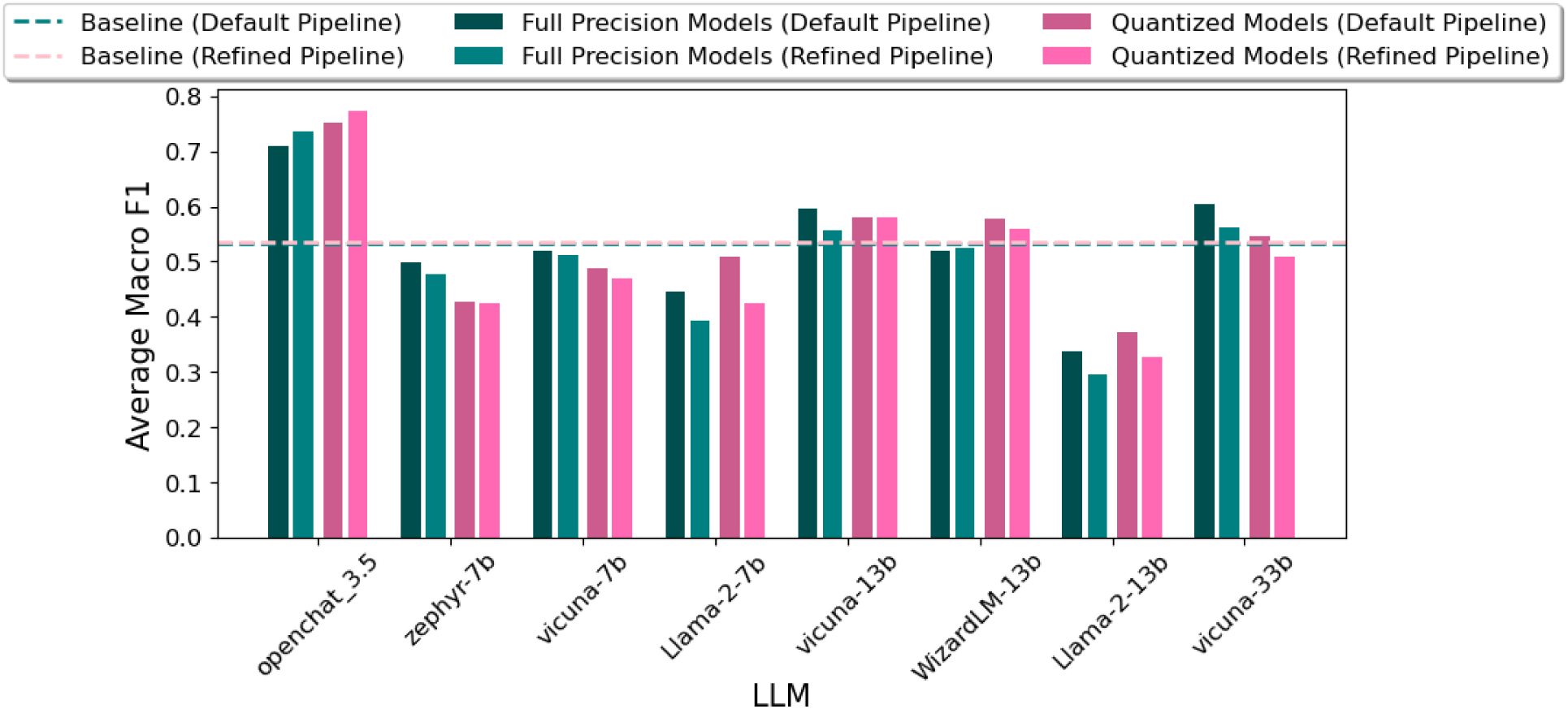
Average Macro F1 score of LLMs.

### Invalid Responses

Figure 4 shows the comparison of the invalid responses between the refined and the default pipeline. It indicates that the refined pipeline greatly reduced the number of invalid responses for each LLM. Some examples of the models’ invalid responses and the corresponding analysis are shown in **Supplementary Table S12**.

**Figure 4.**
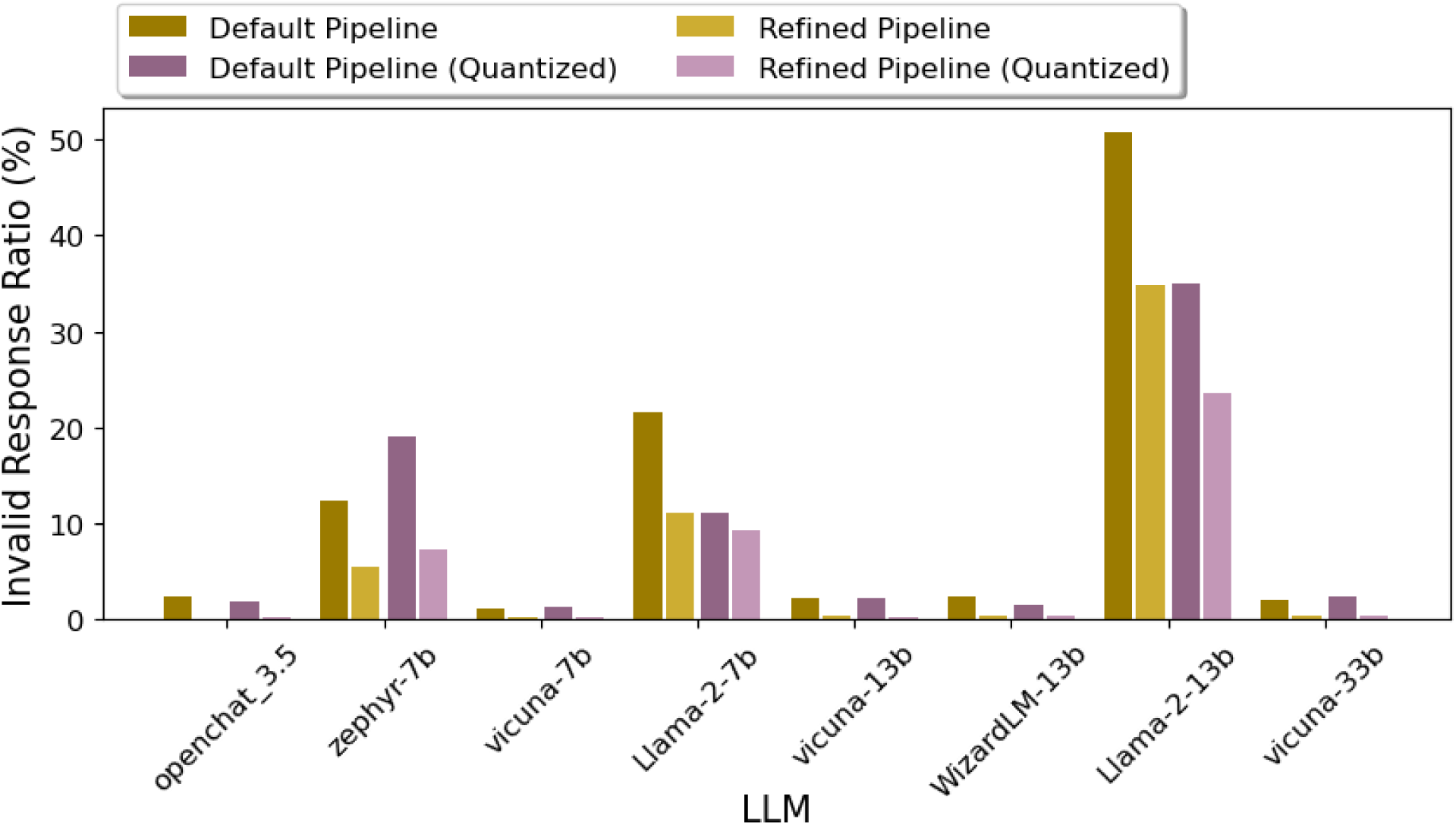
Invalid response rate comparison between the default and the refined pipeline.

## DISCUSSION

In this work, we designed a pipeline that used various open-sourced LLMs to extract patients’ SDoH based on social history documentation in free text EHRs. Using prompt engineering and postprocessing, we demonstrated that some LLMs achieved better performance than the baseline approach of pattern matching on most of the SDoH questions, especially when the answer to the SDoH question was mentioned in the text. We also observed that quantization, which could make these models more applicable in resource limited settings, did not substantially compromise the performance on extraction tasks.

Our study offers some practical learnings regarding using LLMs as scalable extraction tools from free text EHRs. First, thoughtful prompt engineering, as we conducted in our ‘refined’ pipeline, can substantially elevate model performance, and should be routinely considered. Second, manual review of erroneous model responses indicated that in almost all such instances, the text was either controversial or required strong deductive reasoning to infer the correct answer, which is a known challenge for LLMs.[48–50] We also found these cases had a high overlap with the cases reported by the human reviewers as ambiguous or controversial, and the reviewers needed to expand the labeling criteria to reach an agreement on such cases, which showed that such cases are also challenging to humans. Third, we documented clear and large differences between LLMs in this task of information extraction, which underscores the importance of evaluating multiple models when considering specific tasks. Our findings suggesting robust performance of “out of the box” implementation of LLMs without any fine-tuning are complementary to that of a prior report[16], which demonstrated that when resources permit, fine-tuning LLMs for specific extraction tasks including SDOH information can be a viable alternative to further enhance performance.

Our study has important implications. The use of LLM pipelines for generating structured SDoH data from unstructured EHR notes could facilitate numerous downstream applications. One significant area is predictive analytics, where structured SDoH information can be routinely considered alongside structured clinical information to improve risk prediction for various health outcomes. This capability is particularly valuable in managing chronic diseases, where social and environmental factors, such as income and community, play a crucial role. Additionally, structured SDoH data can enhance population health management by enabling healthcare providers to identify and address social disparities in health. In clinical research, the availability of structured SDoH data can refine the selection of study cohorts, improve confounding adjustment in epidemiologic studies, and improve recruitment efforts in clinical trials that aim to oversample socioeconomically disadvantaged patient populations.

The strengths of this work include the automation of the SDoH extraction process with high accuracy. As LLMs avoid the need for model training, this approach is likely scalable and transportable across institutions. Limitations include the complexity and subjectivity involved in the prompt engineering and LLM response postprocessing steps. Further, as most open source LLMs are trained using non-clinical text data, specific training on free-text EHRs maybe needed for extraction of more complex clinical concepts with LLMs and our observations regarding model performance based on general SDoH concepts may not extrapolate to other clinical feature extraction tasks.

## CONCLUSIONS

In conclusion, we demonstrated the feasibility of employing open weight LLMs to extract patients’ SDoH with high accuracy without any additional finetuning. LLMs can offer effective and efficient information extraction from EHR text.

## DECLARATIONS

### Ethics approval and consent to participate

Not applicable.

### Consent for publication

Not applicable.

### Availability of data and materials

The data that support the findings of this study are available from Mass General Brigham (MGB) but restrictions apply to the availability of these data, which were used under license for the current study, and so are not publicly available. Data are however available from the authors upon reasonable request and with permission of Mass General Brigham (MGB).

### Authors’ contributions

BG and VS processed the data, labeled the data, analyzed the data, and drafted the manuscript. ZL contributed the idea of the secondary prompt, which significantly improved the study results. VC and SB contributed the idea of the pipelines in this study. JY and RD proposed the idea of the study, supervised the study, and refined the manuscript. All authors read and approved the final manuscript.

## Data Availability

All data produced in the present study are available from Mass General Brigham (MGB) but restrictions apply to the availability of these data, which were used under license for the current study, and so are not publicly available. Data are however available from the authors upon reasonable request and with permission of Mass General Brigham (MGB).

## LIST OF ABBREVIATIONS

AWQ: Activation-aware Weight Quantization
EHR: Electronic Health Records
LLM: Large Language Model
MGB: Mass General Brigham
NLP: Natural Language Processing
SDoH: Social Determinants of Health

## Acknowledgements

Not applicable.

