## Supplementary for "Scalable information extraction from free text electronic health records using large language models"

**SUPPLEMENTARY TABLE OF CONTENT**

**Annotation Guide**

- Social Determinants of Health (SDoH) annotation guidelines

**Tables**

- Table S1: Distribution of the SDoH Question Choices
- Table S2: LLM Details
- Table S3: The Matching Pattern for the Baseline
- Table S4: Prompt Used in the Pipeline
  - Explanation of Table S4 and Transition to Table S5
- Table S5: [HEADER] Values in Terms of SDoH Questions
- Table S6(a): Model Accuracy_overall_ on Each SDoH Question (Refined Pipeline)
- Table S6(b): Model Accuracy_overall_ on Each SDoH Question (Default Pipeline)
- Table S7(a): Model Accuracy_overall_ Difference (Compared to the Baseline) on Each SDoH Question (Refined Pipeline)
- Table S7(b): Model Accuracy_overall_ Difference (Compared to the Baseline) on Each SDoH Question (Default Pipeline)
- Table S8(a): Model Accuracy_mentioned_ on Each SDoH Question (Refined Pipeline)
- Table S8(b): Model Accuracy_mentioned_ on Each SDoH Question (Default Pipeline)
- Table S9(a): Model Accuracy_mentioned_ Difference (Compared to the Baseline) on Each SDoH Question (Refined Pipeline)
- Table S9(b): Model Accuracy_mentioned_ Difference (Compared to the Baseline) on Each SDoH Question (Default Pipeline)
- Table S10(a): Model Accuracy_non-mentioned_ on Each SDoH Question (Refined Pipeline)
- Table S10(b): Model Accuracy_non-mentioned_ on Each SDoH Question (Default Pipeline)
- Table S11(a): Model Accuracy_non-mentioned_ Difference (Compared to the Baseline) on Each SDoH Question (Refined Pipeline)
- Table S11(b): Model Accuracy_non-mentioned_ Difference (Compared to the Baseline) on Each SDoH Question (Default Pipeline)
- Table S12: Examples of Invalid Responses of Each LLM on the Refined Pipeline (if applicable)

**Social Determinants of Health (SDoH)**

**-** **annotation guidelines-**

Annotation Plan

1)	*Labeling:* each annotator will separately review 100 patient observation notes and manually answer 9 questions related to the 9 social determinants of health

2)	*Initial Validation Phase:* the 2 annotators will collaborative validate their labels for the 100 patient observation notes and resolve any discrepancies to ensure accuracy and agreement of the annotations

EHR data: SDoH annotation involves manual examination of patient observation notes.

Annotation style: Patient observation notes are obtained from the Mass General Brigham Health system. 100 initial patients are randomly selected to create a manually annotated dataset of SDoH which serves as our ground truth for downstream model evaluation. These manual SDoH annotations will be added by reviewing and labeling patient observation notes on the 9 social determinant categories in the labeling phase. Two reviewers will separately review and labels the same patient data and then collaboratively validate patient labels to ensure accuracy and agreement of the annotations in the validation phase.

Annotation types & categories:

- Patient marital status:
  - Single for marital status if:
    - The patient is single (i.e., no spouse or partner)

OR

§ The patient has a boyfriend and/or girlfriend

OR

§ The patient has a long-term partner but is not legally married

- Widowed for marital status if the patient is widowed
- Divorced for marital status if the patient is divorced or separated from their spouse.
- Not mentioned for marital status if:
  - The text does not include any information on patient’s marital status or relationships

OR

- The text states that the patient was not asked about marital status or relationships during visit
- Patient number of children:
  - 0 for number of children if:
    - The patient has no children, including biological, adopted, and step children
  - 1 for number of children if:
    - The patient has 1 child, including biological, adopted, and step children
  - 2 for number of children if:
    - The patient has 2 children, including biological, adopted, and step children
  - 3 for number of children if:
    - The patient has 3 children, including biological, adopted, and step-children
  - 4 for number of children if:
    - The patient has 4 children, including biological, adopted, and step-children
  - 5 or more for number of children if:
    - The patient has 5 or more children, including biological, adopted, and step-children
  - Not mentioned for number of children if:
    - The text does not contain any information related to the patient’s children

OR

- The text states that the patient was not asked about children during the visit
- Patient current tobacco use:
  - Yes for current tobacco use if:
    - The patient currently smokes cigarettes or cigars

OR

- The patient currently uses any form of smokeless tobacco
- No for current tobacco use if:
  - The patient has never used tobacco

OR

- The patient has a history of tobacco use but has quit

OR

- The text mentions past tobacco use but not current tobacco use
- Not mentioned for current tobacco use if:
  - The text does not contain any information related to the patient’s tobacco use

OR

- The text states that the patient was not asked about their tobacco use
- Patient current alcohol use:
  - Yes for current alcohol use if:
    - The patient currently consumes alcohol regularly

OR

- The patient currently consumes alcohol rarely
- No for current alcohol use if:
  - The patient has never consumed alcohol

OR

- The patient has a history of alcohol use but the text does not mention current alcohol use

OR

- The patient is sober from alcohol
- Not mentioned for current alcohol use if:
  - The text does not contain any information related to the patient’s alcohol use

OR

- The text states that the patient was not asked about their alcohol use during visit
- Patient current illicit drug use:
  - Yes for current illicit drug use if:
    - The patient currently uses illicit drugs (e.g., marijuana, cocaine, etc.)
  - No for current illicit drug use if:
    - Patient has never used drugs

OR

- Patient has a history of drug use but no mention of current drug use

OR

- Patient is sober from drugs
- Not mentioned for current illicit drug use if:
  - The text does not contain any information related to the patient’s drug use

OR

- The text states that the patient was not asked about their drug use during visit
- Patient social isolation:
  - Yes for living alone if it is explicitly mentioned in the text that the patient currently lives alone
  - No for living alone if:
    - The patient lives with a family member (i.e., husband/wife/kids)

OR

§ The patient lives with a significant other (i.e., boyfriend/girlfriend)

OR

§ The patient lives with a friend

o	Not mentioned for living alone if:

§ Text does not mention patient’s living situation

OR

§ Text mentions patient has a spouse and/or kids but does not mention living arrangements

OR

§ Text states that patient’s living situation was not asked during visit

·	Patient employment status:

o	Employed for employment status if:

§ Patient currently works full-time

OR

§ Patient currently works part-time

o	Jobless for patient employment status if:

§ Patient is a stay-at-home mom or homemaker

OR

§ Patient temporarily stopped working

OR

§ Patient is unemployed

OR

§ Patient is on disability

o	Retired for patient employment status if:

§ The patient is retired

OR

§ The patient used to work as xxx

OR

§ The patient is a former employee of xxx

o	Not mentioned for patient employment status if:

§ Text does not mention patient’s employment status

OR

§ Text does not mention any patient roles that generate income

OR

§ Text states that patient’s employment status was not asked during visit

·	Patient highest education level:

o	Elementary school for highest education level if:

§ Patient is currently in 1^st^ to 5^th^ grade

OR

§ Patient stopped education between 1^st^ and 5^th^ grade

o	Middle school for highest education level if:

§ Patient is currently in 6^th^ to 8^th^ grade

OR

§ Patient stopped education between 6^th^ and 8^th^ grade

o	High school for highest education level if:

§ Patient is currently in 9^th^ to 12^th^ grade

OR

§ Patient stopped education between 9^th^ and 12^th^ grade

o	College for highest education level if:

§ Patient is currently pursuing or has completed an Associates degree as last form of education.

OR

§ Patient is currently pursuing or has completed a Bachelors degree as last form of education.

OR

§ Patient is currently pursuing a post-bacc or has completed a post-bacc as last form of education.

OR

§ Patient started but did not finish college.

o	Graduate school for highest education level if:

§ Patient is currently pursuing or has completed a Masters degree.

OR

§ Patient is currently pursuing or has completed a Doctoral degree. (i.e., Doctor of Philosophy or Professional Doctoral)

o	Not mentioned for highest education level if:

§ Text does not mention patient’s education

OR

§ Text states that patient’s education was not asked during visit

·	Patient current exercise status:

o	Yes for current exercise status if:

§ Patient currently exercises regularly

OR

§ Patient currently exercises irregularly

o	No for current exercise status if:

§ Patient does not exercise currently

o	In the past for current exercise status if:

§ Patient used to exercise but now does not exercise

OR

§ Patient used to exercise but now cannot due to a physical limitation (i.e., injury, disability)

o	Not mentioned for current exercise status if:

§ Text does not mention patient’s exercise status

OR

§ Text states that patient’s exercise status was not asked during visit

**Table S1. Distribution of the SDoH question choices**

| **SDoH Question** | **SDoH Choices** | **Appearance Count (Validation set, n=100)** | **Appearance Count (Test set, n=100)** |
| --- | --- | --- | --- |
| Q1. What is the patient’s marital status? | Single | 28 | 34 |
|  | Widowed | 10 | 7 |
|  | Divorced | 6 | 9 |
|  | Married | 49 | 41 |
|  | Not mentioned | 7 | 9 |
| Q2. How many children does the patient have? | 0 | 3 | 7 |
|  | 1 | 21 | 22 |
|  | 2 | 16 | 18 |
|  | 3 | 7 | 9 |
|  | 4 | 4 | 0 |
|  | 5 or more | 3 | 4 |
|  | Not mentioned | 46 | 40 |
| Q3. Does the patient currently use tobacco? | Yes | 3 | 1 |
|  | No | 17 | 16 |
|  | Not mentioned | 80 | 83 |
| Q4. Does the patient currently consume alcohol? | Yes | 11 | 10 |
|  | No | 9 | 3 |
|  | Not mentioned | 80 | 87 |
| Q5. Does the patient currently use illicit drugs? | Yes | 0 | 1 |
|  | No | 9 | 7 |
|  | Not mentioned | 91 | 92 |
| Q6. Does the patient live alone? | Yes | 8 | 7 |
|  | No | 48 | 54 |
|  | Not mentioned | 44 | 39 |
| Q7. What is the patient’s employment status? | Employed | 34 | 28 |
|  | Jobless | 12 | 14 |
|  | Retired | 27 | 26 |
|  | Not mentioned | 27 | 32 |
| Q8. What is the patient’s highest education level? | Elementary school | 1 | 2 |
|  | Middle school | 1 | 0 |
|  | High school | 3 | 7 |
|  | College | 4 | 12 |
|  | Graduate school | 4 | 2 |
|  | Not mentioned | 87 | 77 |
| Q9. Does the patient exercise? | Yes | 15 | 10 |
|  | No | 0 | 0 |
|  | In the past | 1 | 0 |
|  | Not mentioned | 84 | 90 |

**Table S2. LLM details**

| **Model Name** | **Parameter Size** | **Hugging Face Link** |
| --- | --- | --- |
| openchat_3.5 | 7B | https://huggingface.co/openchat/openchat_3.5 |
| zephyr-7b-beta | 7B | https://huggingface.co/HuggingFaceH4/zephyr-7b-beta |
| vicuna-7b-v1.5 | 7B | https://huggingface.co/lmsys/vicuna-7b-v1.5 |
| Llama-2-7b-chat-hf | 7B | https://huggingface.co/meta-llama/Llama-2-7b-chat-hf |
| vicuna-13b-v1.5 | 13B | https://huggingface.co/lmsys/vicuna-13b-v1.5 |
| WizardLM-13B-V1.2 | 13B | https://huggingface.co/WizardLM/WizardLM-13B-V1.2 |
| Llama-2-13b-chat-hf | 13B | https://huggingface.co/meta-llama/Llama-2-13b-chat-hf |
| vicuna-33b-v1.3 | 33B | https://huggingface.co/lmsys/vicuna-33b-v1.3 |

**Table S3. The matching pattern for the baseline**

| **SDoH Question** | **Matching Pattern (Case Insensitive)** | **Resulting Choice** |
| --- | --- | --- |
| Q1. What is the patient’s marital status? | married | Married |
|  | widowed | Widowed |
|  | divorced\|separated | Divorced |
|  | single\|no spouse\|partner\|boyfriend\|girlfriend\|long-term partner | Single |
|  | default | Not mentioned |
| Q2. How many children does the patient have? | no children\|0 children | 0 |
|  | 1 child\|1 son\|1 daughter | 1 |
|  | 2 children\|2 son\|2 daughter | 2 |
|  | 3 children\|3 son\|3 daughter | 3 |
|  | 4 children\|4 son\|4 daughter | 4 |
|  | 5 children\|6 children\|7 children\|8 children\|9 children\|10 children | 5 or more |
|  | default | Not mentioned |
| Q3. Does the patient currently use tobacco? | currently smokes\|cigarettes\|cigars\|smokeless tobacco\|tobacco use\|smoke | Yes |
|  | never used tobacco\|quit smoking\|quit tobacco\|quit smoke\|past tobacco use\|never smoke\|no smoke | No |
|  | default | Not mentioned |
| Q4. Does the patient currently consume alcohol? | consumes alcohol\|drinks alcohol\|ETOH | Yes |
|  | never consumed alcohol\|sober\|no ETOH | No |
|  | default | Not mentioned |
| Q5. Does the patient currently use illicit drugs? | illicit drugs\|uses drugs\|cocaine\|marijuana\|substance abuse | Yes |
|  | never used drugs\|sober from drugs\|past drug use\|deny illicit drugs | No |
|  | default | Not mentioned |
| Q6. Does the patient live alone? | alone | Yes |
|  | with husband\|with wife\|with child\|with children\|with son\|with daughter\|with boyfriend\|with girlfriend\|with friend\|with grandparents\|with grandmother\|with grandfather\|with uncle\|with aunt\|with parents\|with father\|with mother\|with dad\|with mom | No |
|  | default | Not mentioned |
| Q7. What is the patient’s employment status? | full-time\|part-time\|employed\|work as\|employ | Employed |
|  | stay-at-home\|at home\|unemployed\|on disability\|homemaker\|not work\|no work\|jobless | Jobless |
|  | retire\|former employee\|worked\|used to work | Retired |
|  | default | Not mentioned |
| Q8. What is the patient’s highest education level? | elementary school\|1st grade\|2nd grade\|3rd grade\|4th grade\|5th grade | Elementary school |
|  | middle school\|6th grade\|7th grade\|8th grade | Middle school |
|  | high school\|9th grade\|10th grade\|11th grade | High school |
|  | college\|Associates\|Bachelors\|BS\|BA\|freshman\|sophomore\|junior\|senior\|post-bacc | College |
|  | graduate school\|grad school\|Masters degree\|MS\|MBA\|MPH\|MENG\|Doctoral degree\|PHD\|MD | Graduate school |
|  | default | Not mentioned |
| Q9. Does the patient exercise? | exercise\|walk\|run\|dance\|gym\|treadmill\|yoga | Yes |
|  | does not exercise\|never exercise\|not exercising | No |
|  | used to exercise\|used to walk | In the past |
|  | default | Not mentioned |

**Table S4. Prompt used in the pipeline**

| **Prompt Name** | **Prompt Content** |
| --- | --- |
| Default prompt | *USER: Given the following text about a patient's social documentation.*  *[TEXT]*  *Please answer the question: [QUESTION]*  *ASSISTANT:* |
| Premise prompt for Q2 | *USER: Given the following text about a patient's social documentation.*  *[TEXT]*  *Please answer the question: Does the text mention the patient's children (including no children)?*  *Your answer must be one of the following:*  *- Yes*  *- No*  *ASSISTANT:* |
| Special prompt for Q6 | *USER: Given the following text about a patient's social documentation.*  *[TEXT]*  *Please answer the question: [QUESTION]*  *Please DO NOT infer the answer according to the text. ONLY give your answers based on the ACTUAL text.*  *Your answer should be "Yes" IF AND ONLY IF the text EXPLICITLY mentions that the patient lives alone.*  *Your answer should be "No" IF AND ONLY IF the text EXPLICITLY mentions that the patient lives with someone.*  *Otherwise, your answer MUST be "Not mentioned".*  *ASSISTANT:* |
| Special prompt for Q7 | *USER: Given the following text about a patient's social documentation.*  *[TEXT]*  *Please answer the question: [QUESTION]*  *Your answer should be "Employed" if the text mentions that the patient has a full-time or part-time job (even if the patient retired or was jobless before).*  *Your answer should be "Jobless" if the text mentions that the patient is not working, is a homemaker, or is on disability.*  *Your answer should be "Retired" if the text mentions that the patient is retired, is a former employee or used to be an employee.*  *ASSISTANT:* |
| Default secondary prompt | *USER: Suppose that [HEADER] [FIRST PROMPT MODEL RESPONSE].*  *[QUESTION]*  *ASSISTANT:* |
| Secondary prompt for Q7 | *USER: Given the following text about a patient's social documentation.*  *[FIRST PROMPT MODEL RESPONSE]*  *Please answer the question: Does the text mention the patient's occupation?*  *Your answer must be one of the following:*  *- Yes*  *- No*  *ASSISTANT:* |

***Explanation of Table S4 and Transition to Table S5***

The default prompt was the basic prompt that asked the model to respond using one of the choices of the SDoH questions and did not involve any prompt engineering techniques. The premise prompt was a prompt that provided pre-specified input text to the model before asking the actual SDoH question. The purpose of the premise prompt was to reduce model hallucination. It queried the model if the target information was present in the text before asking the actual SDoH question and the answer could only be either “Yes” or “No”. If the model responded with “No”, the response to the actual SDoH question would be defined to be “Not mentioned” without asking the model the actual SDoH question. If the model responded with “Yes”, then the actual SDoH question was asked. According to our experiments with the validation set, we only applied such premise prompts to Q2. The special prompt was a prompt with prompt engineering, which was built for specific questions to improve the model accuracy. Based on our experiments on the validation set, such prompts were applied to Q6 and Q7. The secondary prompt was a prompt that was used to reduce the number of invalid responses. Such a prompt was used when the initial model response could not be mapped to a single choice by the post-processing procedure. Secondary prompts were used for all 9 SDoH questions. The special case of the secondary prompts was the “Secondary prompt for Q7”, which was a prompt that was used to enforce the model response to be “Employed” if the initial model response was “Not mentioned”, but the patient’s occupation was shown in the text. The intuition of such a prompt was that for many models, they responded “Not mentioned” to Q7 if the input text only mentioned the patient’s occupation but did not mention whether the occupation is a current or previous one. However, according to the labeling criteria, such condition should be considered as “Employed”. As a result, the “Secondary prompt for Q7” was used to enforce this assumption.

The “[TEXT]” is where we substitute for the patients’ social documentation, and the “[QUESTION]” is where we substitute for the actual SDoH questions. The “[FIRST PROMPT MODEL RESPONSE]” is where we substitute for the LLM’s response to this question generated on the primary prompt. The “[HEADER]” is where we substitute for a string that has the value shown in **Table S5** based on different SDoH questions. For each prompt mentioned in **Table S4**, we added a two-sentence header before the context of the prompt which read as follows.

*A chat between a curious user and an artificial intelligence assistant. The assistant gives helpful, detailed, and polite answers to the user's questions.*

The two sentences above originated from the LMSYS Org, the team that created the Vicuna LLM, and was used during the pre-training phase of the LLM. The header was later adopted by the WizardLM (Can et al, 2023) LLM during its pre-training phase. During our experiment, we noticed that the headers are essential for the Vicuna and WizardLM LLMs to give reasonable responses. For other LLMs, since they do not have a prompting template specified, we applied the same header on them for consistency.

**Table S5. [HEADER] values in terms of SDoH questions**

| **SDoH Question** | **[HEADER] Value** |
| --- | --- |
| Q1. What is the patient’s marital status? | the patient’s marital status information is: |
| Q2. How many children does the patient have? | the patient's children information is: |
| Q3. Does the patient currently use tobacco? | the patient's smoking or tobacco usage information is: |
| Q4. Does the patient currently consume alcohol? | the patient's drinking or alcohol usage information is: |
| Q5. Does the patient currently use illicit drugs? | the patient's drug usage information is: |
| Q6. Does the patient live alone? | the patient's living information is: |
| Q7. What is the patient’s employment status? | the patient's employment information is: |
| Q8. What is the patient’s highest education level? | the patient's education information is: |
| Q9. Does the patient exercise? | the patient's exercising information is: |

**Table S6(a). Model Accuracy_overall_ on each SDoH question (Refined pipeline)**

| **Model** | **Q1** | **Q2** | **Q3** | **Q4** | **Q5** | **Q6** | **Q7** | **Q8** | **Q9** |
| --- | --- | --- | --- | --- | --- | --- | --- | --- | --- |
| openchat_3.5 | 96.0 (95.96, 96.04) | 83.0 (82.92, 83.08) | 96.0 (95.96, 96.04) | 93.0 (92.95, 93.05) | 97.0 (96.97, 97.03) | 85.0 (84.93, 85.07) | 78.0 (77.92, 78.08) | 94.0 (93.95, 94.05) | 97.0 (96.97, 97.03) |
| openchat_3.5-w4-g128 | 96.0 (95.96, 96.04) | 80.0 (79.92, 80.08) | 96.0 (95.96, 96.04) | 95.0 (94.96, 95.04) | 96.0 (95.96, 96.04) | 86.0 (85.93, 86.07) | 79.0 (78.92, 79.08) | 92.0 (91.95, 92.05) | 97.0 (96.97, 97.03) |
| zephyr-7b-beta | 72.0 (71.91, 72.09) | 60.0 (59.9, 60.1) | 82.0 (81.92, 82.08) | 87.0 (86.93, 87.07) | 79.0 (78.92, 79.08) | 42.0 (41.9, 42.1) | 62.0 (61.9, 62.1) | 70.0 (69.91, 70.09) | 88.0 (87.93, 88.07) |
| zephyr-7b-beta-w4-g128 | 75.0 (74.91, 75.09) | 55.0 (54.9, 55.1) | 61.0 (60.9, 61.1) | 62.0 (61.9, 62.1) | 69.0 (68.91, 69.09) | 47.0 (46.9, 47.1) | 50.0 (49.9, 50.1) | 62.0 (61.9, 62.1) | 90.0 (89.94, 90.06) |
| vicuna-7b-v1.5 | 83.0 (82.92, 83.08) | 70.0 (69.91, 70.09) | 86.0 (85.93, 86.07) | 89.0 (88.94, 89.06) | 92.0 (91.95, 92.05) | 42.0 (41.9, 42.1) | 63.0 (62.9, 63.1) | 64.0 (63.9, 64.1) | 60.0 (59.9, 60.1) |
| vicuna-7b-v1.5-w4-g128 | 79.0 (78.92, 79.08) | 63.0 (62.9, 63.1) | 85.0 (84.93, 85.07) | 87.0 (86.93, 87.07) | 93.0 (92.95, 93.05) | 40.0 (39.9, 40.1) | 65.0 (64.9, 65.1) | 80.0 (79.92, 80.08) | 79.0 (78.92, 79.08) |
| Llama-2-7b-chat-hf | 60.0 (59.9, 60.1) | 58.0 (57.9, 58.1) | 73.0 (72.91, 73.09) | 79.0 (78.92, 79.08) | 48.0 (47.9, 48.1) | 36.0 (35.9, 36.1) | 23.0 (22.92, 23.08) | 35.0 (34.9, 35.1) | 82.0 (81.92, 82.08) |
| Llama-2-7b-chat-hf-w4-g128 | 86.0 (85.93, 86.07) | 55.0 (54.9, 55.1) | 63.0 (62.9, 63.1) | 80.0 (79.92, 80.08) | 26.0 (25.91, 26.09) | 35.0 (34.9, 35.1) | 31.0 (30.91, 31.09) | 40.0 (39.9, 40.1) | 68.0 (67.91, 68.09) |
| vicuna-13b-v1.5 | 95.0 (94.96, 95.04) | 78.0 (77.92, 78.08) | 94.0 (93.95, 94.05) | 90.0 (89.94, 90.06) | 91.0 (90.94, 91.06) | 63.0 (62.9, 63.1) | 33.0 (32.91, 33.09) | 84.0 (83.93, 84.07) | 89.0 (88.94, 89.06) |
| vicuna-13b-v1.5-w4-g128 | 92.0 (91.95, 92.05) | 73.0 (72.91, 73.09) | 96.0 (95.96, 96.04) | 89.0 (88.94, 89.06) | 94.0 (93.95, 94.05) | 55.0 (54.9, 55.1) | 43.0 (42.9, 43.1) | 88.0 (87.93, 88.07) | 90.0 (89.94, 90.06) |
| WizardLM-13B-V1.2 | 89.0 (88.94, 89.06) | 77.0 (76.92, 77.08) | 79.0 (78.92, 79.08) | 77.0 (76.92, 77.08) | 80.0 (79.92, 80.08) | 51.0 (50.9, 51.1) | 63.0 (62.9, 63.1) | 73.0 (72.91, 73.09) | 79.0 (78.92, 79.08) |
| WizardLM-13B-V1.2-w4-g128 | 90.0 (89.94, 90.06) | 76.0 (75.91, 76.09) | 91.0 (90.94, 91.06) | 90.0 (89.94, 90.06) | 93.0 (92.95, 93.05) | 62.0 (61.9, 62.1) | 56.0 (55.9, 56.1) | 74.0 (73.91, 74.09) | 88.0 (87.93, 88.07) |
| Llama-2-13b-chat-hf | 36.0 (35.9, 36.1) | 33.0 (32.91, 33.09) | 45.0 (44.9, 45.1) | 27.0 (26.91, 27.09) | 47.0 (46.9, 47.1) | 26.0 (25.91, 26.09) | 24.0 (23.91, 24.09) | 40.0 (39.9, 40.1) | 68.0 (67.91, 68.09) |
| Llama-2-13b-chat-hf-w4-g128 | 53.0 (52.9, 53.1) | 30.0 (29.91, 30.09) | 68.0 (67.91, 68.09) | 56.0 (55.9, 56.1) | 65.0 (64.9, 65.1) | 27.0 (26.91, 27.09) | 16.0 (15.93, 16.07) | 51.0 (50.9, 51.1) | 86.0 (85.93, 86.07) |
| vicuna-33b-v1.3 | 61.0 (60.9, 61.1) | 78.0 (77.92, 78.08) | 90.0 (89.94, 90.06) | 94.0 (93.95, 94.05) | 83.0 (82.92, 83.08) | 42.0 (41.9, 42.1) | 56.0 (55.9, 56.1) | 77.0 (76.92, 77.08) | 94.0 (93.95, 94.05) |
| vicuna-33b-v1.3-w4-g128 | 59.0 (58.9, 59.1) | 79.0 (78.92, 79.08) | 88.0 (87.93, 88.07) | 93.0 (92.95, 93.05) | 85.0 (84.93, 85.07) | 41.0 (40.9, 41.1) | 52.0 (51.9, 52.1) | 79.0 (78.92, 79.08) | 93.0 (92.95, 93.05) |
| baseline | 92.0 (91.95, 92.05) | 51.0 (50.9, 51.1) | 83.0 (82.92, 83.08) | 92.0 (91.95, 92.05) | 93.0 (92.95, 93.05) | 68.0 (67.91, 68.09) | 42.0 (41.9, 42.1) | 82.0 (81.92, 82.08) | 93.0 (92.95, 93.05) |

**Table S6(b). Model Accuracy_overall_ on each SDoH question (Default pipeline)**

| **Model** | **Q1** | **Q2** | **Q3** | **Q4** | **Q5** | **Q6** | **Q7** | **Q8** | **Q9** |
| --- | --- | --- | --- | --- | --- | --- | --- | --- | --- |
| openchat_3.5 | 96.0 (95.96, 96.04) | 72.0 (71.91, 72.09) | 96.0 (95.96, 96.04) | 93.0 (92.95, 93.05) | 96.0 (95.96, 96.04) | 76.0 (75.91, 76.09) | 74.0 (73.91, 74.09) | 92.0 (91.95, 92.05) | 97.0 (96.97, 97.03) |
| openchat_3.5-w4-g128 | 96.0 (95.96, 96.04) | 69.0 (68.91, 69.09) | 96.0 (95.96, 96.04) | 95.0 (94.96, 95.04) | 95.0 (94.96, 95.04) | 75.0 (74.91, 75.09) | 72.0 (71.91, 72.09) | 91.0 (90.94, 91.06) | 97.0 (96.97, 97.03) |
| zephyr-7b-beta | 71.0 (70.91, 71.09) | 60.0 (59.9, 60.1) | 79.0 (78.92, 79.08) | 84.0 (83.93, 84.07) | 74.0 (73.91, 74.09) | 64.0 (63.9, 64.1) | 57.0 (56.9, 57.1) | 56.0 (55.9, 56.1) | 86.0 (85.93, 86.07) |
| zephyr-7b-beta-w4-g128 | 75.0 (74.91, 75.09) | 51.0 (50.9, 51.1) | 54.0 (53.9, 54.1) | 60.0 (59.9, 60.1) | 64.0 (63.9, 64.1) | 42.0 (41.9, 42.1) | 56.0 (55.9, 56.1) | 43.0 (42.9, 43.1) | 80.0 (79.92, 80.08) |
| vicuna-7b-v1.5 | 82.0 (81.92, 82.08) | 47.0 (46.9, 47.1) | 85.0 (84.93, 85.07) | 88.0 (87.93, 88.07) | 91.0 (90.94, 91.06) | 59.0 (58.9, 59.1) | 49.0 (48.9, 49.1) | 63.0 (62.9, 63.1) | 60.0 (59.9, 60.1) |
| vicuna-7b-v1.5-w4-g128 | 78.0 (77.92, 78.08) | 45.0 (44.9, 45.1) | 84.0 (83.93, 84.07) | 86.0 (85.93, 86.07) | 92.0 (91.95, 92.05) | 59.0 (58.9, 59.1) | 46.0 (45.9, 46.1) | 79.0 (78.92, 79.08) | 78.0 (77.92, 78.08) |
| Llama-2-7b-chat-hf | 61.0 (60.9, 61.1) | 39.0 (38.9, 39.1) | 60.0 (59.9, 60.1) | 71.0 (70.91, 71.09) | 45.0 (44.9, 45.1) | 54.0 (53.9, 54.1) | 46.0 (45.9, 46.1) | 28.0 (27.91, 28.09) | 82.0 (81.92, 82.08) |
| Llama-2-7b-chat-hf-w4-g128 | 84.0 (83.93, 84.07) | 52.0 (51.9, 52.1) | 61.0 (60.9, 61.1) | 79.0 (78.92, 79.08) | 24.0 (23.91, 24.09) | 60.0 (59.9, 60.1) | 56.0 (55.9, 56.1) | 38.0 (37.9, 38.1) | 68.0 (67.91, 68.09) |
| vicuna-13b-v1.5 | 94.0 (93.95, 94.05) | 47.0 (46.9, 47.1) | 93.0 (92.95, 93.05) | 89.0 (88.94, 89.06) | 90.0 (89.94, 90.06) | 80.0 (79.92, 80.08) | 75.0 (74.91, 75.09) | 80.0 (79.92, 80.08) | 88.0 (87.93, 88.07) |
| vicuna-13b-v1.5-w4-g128 | 92.0 (91.95, 92.05) | 46.0 (45.9, 46.1) | 94.0 (93.95, 94.05) | 88.0 (87.93, 88.07) | 93.0 (92.95, 93.05) | 71.0 (70.91, 71.09) | 73.0 (72.91, 73.09) | 84.0 (83.93, 84.07) | 89.0 (88.94, 89.06) |
| WizardLM-13B-V1.2 | 89.0 (88.94, 89.06) | 68.0 (67.91, 68.09) | 79.0 (78.92, 79.08) | 76.0 (75.91, 76.09) | 79.0 (78.92, 79.08) | 58.0 (57.9, 58.1) | 55.0 (54.9, 55.1) | 66.0 (65.9, 66.1) | 78.0 (77.92, 78.08) |
| WizardLM-13B-V1.2-w4-g128 | 90.0 (89.94, 90.06) | 67.0 (66.91, 67.09) | 91.0 (90.94, 91.06) | 90.0 (89.94, 90.06) | 93.0 (92.95, 93.05) | 70.0 (69.91, 70.09) | 66.0 (65.9, 66.1) | 71.0 (70.91, 71.09) | 88.0 (87.93, 88.07) |
| Llama-2-13b-chat-hf | 27.0 (26.91, 27.09) | 17.0 (16.92, 17.08) | 32.0 (31.91, 32.09) | 30.0 (29.91, 30.09) | 39.0 (38.9, 39.1) | 30.0 (29.91, 30.09) | 24.0 (23.91, 24.09) | 39.0 (38.9, 39.1) | 56.0 (55.9, 56.1) |
| Llama-2-13b-chat-hf-w4-g128 | 51.0 (50.9, 51.1) | 12.0 (11.93, 12.07) | 67.0 (66.91, 67.09) | 56.0 (55.9, 56.1) | 51.0 (50.9, 51.1) | 36.0 (35.9, 36.1) | 37.0 (36.9, 37.1) | 41.0 (40.9, 41.1) | 72.0 (71.91, 72.09) |
| vicuna-33b-v1.3 | 61.0 (60.9, 61.1) | 56.0 (55.9, 56.1) | 90.0 (89.94, 90.06) | 93.0 (92.95, 93.05) | 83.0 (82.92, 83.08) | 80.0 (79.92, 80.08) | 66.0 (65.9, 66.1) | 75.0 (74.91, 75.09) | 94.0 (93.95, 94.05) |
| vicuna-33b-v1.3-w4-g128 | 59.0 (58.9, 59.1) | 64.0 (63.9, 64.1) | 88.0 (87.93, 88.07) | 93.0 (92.95, 93.05) | 84.0 (83.93, 84.07) | 68.0 (67.91, 68.09) | 65.0 (64.9, 65.1) | 77.0 (76.92, 77.08) | 92.0 (91.95, 92.05) |
| baseline | 92.0 (91.95, 92.05) | 51.0 (50.9, 51.1) | 83.0 (82.92, 83.08) | 92.0 (91.95, 92.05) | 93.0 (92.95, 93.05) | 65.0 (64.9, 65.1) | 42.0 (41.9, 42.1) | 82.0 (81.92, 82.08) | 93.0 (92.95, 93.05) |

**Table S7 (a). Model Accuracy_overall_ difference (compared to the baseline) on each SDoH question (Refined pipeline)**

| **Model** | **Q1** | **Q2** | **Q3** | **Q4** | **Q5** | **Q6** | **Q7** | **Q8** | **Q9** |
| --- | --- | --- | --- | --- | --- | --- | --- | --- | --- |
| openchat_3.5 | 4.0 (3.93, 4.07) | 32.0 (31.87, 32.13) | 13.0 (12.92, 13.08) | 1.0 (0.93, 1.07) | 4.0 (3.94, 4.06) | 17.0 (16.88, 17.12) | 36.0 (35.87, 36.13) | 12.0 (11.91, 12.09) | 4.0 (3.94, 4.06) |
| openchat_3.5-w4-g128 | 4.0 (3.93, 4.07) | 29.0 (28.87, 29.13) | 13.0 (12.92, 13.08) | 3.0 (2.93, 3.07) | 3.0 (2.94, 3.06) | 18.0 (17.88, 18.12) | 37.0 (36.87, 37.13) | 10.0 (9.91, 10.09) | 4.0 (3.94, 4.06) |
| zephyr-7b-beta | -20.0 (-20.11, -19.89) | 9.0 (8.86, 9.14) | -1.0 (-1.11, -0.89) | -5.0 (-5.09, -4.91) | -14.0 (-14.1, -13.9) | -26.0 (-26.14, -25.86) | 20.0 (19.86, 20.14) | -12.0 (-12.12, -11.88) | -5.0 (-5.08, -4.92) |
| zephyr-7b-beta-w4-g128 | -17.0 (-17.1, -16.9) | 4.0 (3.86, 4.14) | -22.0 (-22.12, -21.88) | -30.0 (-30.11, -29.89) | -24.0 (-24.11, -23.89) | -21.0 (-21.14, -20.86) | 8.0 (7.86, 8.14) | -20.0 (-20.12, -19.88) | -3.0 (-3.08, -2.92) |
| vicuna-7b-v1.5 | -9.0 (-9.09, -8.91) | 19.0 (18.86, 19.14) | 3.0 (2.9, 3.1) | -3.0 (-3.08, -2.92) | -1.0 (-1.07, -0.93) | -26.0 (-26.14, -25.86) | 21.0 (20.86, 21.14) | -18.0 (-18.12, -17.88) | -33.0 (-33.11, -32.89) |
| vicuna-7b-v1.5-w4-g128 | -13.0 (-13.1, -12.9) | 12.0 (11.86, 12.14) | 2.0 (1.9, 2.1) | -5.0 (-5.09, -4.91) | 0.0 (-0.07, 0.07) | -28.0 (-28.14, -27.86) | 23.0 (22.86, 23.14) | -2.0 (-2.11, -1.89) | -14.0 (-14.1, -13.9) |
| Llama-2-7b-chat-hf | -32.0 (-32.11, -31.89) | 7.0 (6.86, 7.14) | -10.0 (-10.12, -9.88) | -13.0 (-13.1, -12.9) | -45.0 (-45.11, -44.89) | -32.0 (-32.13, -31.87) | -19.0 (-19.13, -18.87) | -47.0 (-47.12, -46.88) | -11.0 (-11.09, -10.91) |
| Llama-2-7b-chat-hf-w4-g128 | -6.0 (-6.09, -5.91) | 4.0 (3.86, 4.14) | -20.0 (-20.12, -19.88) | -12.0 (-12.1, -11.9) | -67.0 (-67.1, -66.9) | -33.0 (-33.13, -32.87) | -11.0 (-11.14, -10.86) | -42.0 (-42.12, -41.88) | -25.0 (-25.11, -24.89) |
| vicuna-13b-v1.5 | 3.0 (2.93, 3.07) | 27.0 (26.87, 27.13) | 11.0 (10.91, 11.09) | -2.0 (-2.08, -1.92) | -2.0 (-2.08, -1.92) | -5.0 (-5.13, -4.87) | -9.0 (-9.14, -8.86) | 2.0 (1.89, 2.11) | -4.0 (-4.08, -3.92) |
| vicuna-13b-v1.5-w4-g128 | 0.0 (-0.08, 0.08) | 22.0 (21.87, 22.13) | 13.0 (12.92, 13.08) | -3.0 (-3.08, -2.92) | 1.0 (0.93, 1.07) | -13.0 (-13.14, -12.86) | 1.0 (0.86, 1.14) | 6.0 (5.9, 6.1) | -3.0 (-3.08, -2.92) |
| WizardLM-13B-V1.2 | -3.0 (-3.08, -2.92) | 26.0 (25.87, 26.13) | -4.0 (-4.11, -3.89) | -15.0 (-15.1, -14.9) | -13.0 (-13.1, -12.9) | -17.0 (-17.14, -16.86) | 21.0 (20.86, 21.14) | -9.0 (-9.12, -8.88) | -14.0 (-14.1, -13.9) |
| WizardLM-13B-V1.2-w4-g128 | -2.0 (-2.08, -1.92) | 25.0 (24.87, 25.13) | 8.0 (7.91, 8.09) | -2.0 (-2.08, -1.92) | 0.0 (-0.07, 0.07) | -6.0 (-6.13, -5.87) | 14.0 (13.86, 14.14) | -8.0 (-8.12, -7.88) | -5.0 (-5.08, -4.92) |
| Llama-2-13b-chat-hf | -56.0 (-56.11, -55.89) | -18.0 (-18.14, -17.86) | -38.0 (-38.12, -37.88) | -65.0 (-65.1, -64.9) | -46.0 (-46.11, -45.89) | -42.0 (-42.13, -41.87) | -18.0 (-18.13, -17.87) | -42.0 (-42.12, -41.88) | -25.0 (-25.11, -24.89) |
| Llama-2-13b-chat-hf-w4-g128 | -39.0 (-39.11, -38.89) | -21.0 (-21.14, -20.86) | -15.0 (-15.12, -14.88) | -36.0 (-36.11, -35.89) | -28.0 (-28.11, -27.89) | -41.0 (-41.13, -40.87) | -26.0 (-26.12, -25.88) | -31.0 (-31.13, -30.87) | -7.0 (-7.09, -6.91) |
| vicuna-33b-v1.3 | -31.0 (-31.11, -30.89) | 27.0 (26.87, 27.13) | 7.0 (6.9, 7.1) | 2.0 (1.93, 2.07) | -10.0 (-10.09, -9.91) | -26.0 (-26.14, -25.86) | 14.0 (13.86, 14.14) | -5.0 (-5.11, -4.89) | 1.0 (0.93, 1.07) |
| vicuna-33b-v1.3-w4-g128 | -33.0 (-33.11, -32.89) | 28.0 (27.87, 28.13) | 5.0 (4.9, 5.1) | 1.0 (0.93, 1.07) | -8.0 (-8.09, -7.91) | -27.0 (-27.14, -26.86) | 10.0 (9.86, 10.14) | -3.0 (-3.11, -2.89) | 0.0 (-0.07, 0.07) |

**Table S7 (b). Model Accuracy_overall_ difference (compared to the baseline) on each SDoH question (Default pipeline)**

| **Model** | **Q1** | **Q2** | **Q3** | **Q4** | **Q5** | **Q6** | **Q7** | **Q8** | **Q9** |
| --- | --- | --- | --- | --- | --- | --- | --- | --- | --- |
| openchat_3.5 | 4.0 (3.93, 4.07) | 21.0 (20.87, 21.13) | 13.0 (12.92, 13.08) | 1.0 (0.93, 1.07) | 3.0 (2.94, 3.06) | 11.0 (10.87, 11.13) | 32.0 (31.87, 32.13) | 10.0 (9.91, 10.09) | 4.0 (3.94, 4.06) |
| openchat_3.5-w4-g128 | 4.0 (3.93, 4.07) | 18.0 (17.86, 18.14) | 13.0 (12.92, 13.08) | 3.0 (2.93, 3.07) | 2.0 (1.93, 2.07) | 10.0 (9.87, 10.13) | 30.0 (29.87, 30.13) | 9.0 (8.9, 9.1) | 4.0 (3.94, 4.06) |
| zephyr-7b-beta | -21.0 (-21.11, -20.89) | 9.0 (8.86, 9.14) | -4.0 (-4.11, -3.89) | -8.0 (-8.09, -7.91) | -19.0 (-19.1, -18.9) | -1.0 (-1.14, -0.86) | 15.0 (14.86, 15.14) | -26.0 (-26.13, -25.87) | -7.0 (-7.09, -6.91) |
| zephyr-7b-beta-w4-g128 | -17.0 (-17.1, -16.9) | 0.0 (-0.14, 0.14) | -29.0 (-29.13, -28.87) | -32.0 (-32.11, -31.89) | -29.0 (-29.11, -28.89) | -23.0 (-23.14, -22.86) | 14.0 (13.86, 14.14) | -39.0 (-39.13, -38.87) | -13.0 (-13.1, -12.9) |
| vicuna-7b-v1.5 | -10.0 (-10.09, -9.91) | -4.0 (-4.14, -3.86) | 2.0 (1.9, 2.1) | -4.0 (-4.08, -3.92) | -2.0 (-2.08, -1.92) | -6.0 (-6.14, -5.86) | 7.0 (6.86, 7.14) | -19.0 (-19.12, -18.88) | -33.0 (-33.11, -32.89) |
| vicuna-7b-v1.5-w4-g128 | -14.0 (-14.1, -13.9) | -6.0 (-6.14, -5.86) | 1.0 (0.89, 1.11) | -6.0 (-6.09, -5.91) | -1.0 (-1.07, -0.93) | -6.0 (-6.14, -5.86) | 4.0 (3.86, 4.14) | -3.0 (-3.11, -2.89) | -15.0 (-15.1, -14.9) |
| Llama-2-7b-chat-hf | -31.0 (-31.11, -30.89) | -12.0 (-12.14, -11.86) | -23.0 (-23.12, -22.88) | -21.0 (-21.11, -20.89) | -48.0 (-48.11, -47.89) | -11.0 (-11.14, -10.86) | 4.0 (3.86, 4.14) | -54.0 (-54.12, -53.88) | -11.0 (-11.09, -10.91) |
| Llama-2-7b-chat-hf-w4-g128 | -8.0 (-8.09, -7.91) | 1.0 (0.86, 1.14) | -22.0 (-22.12, -21.88) | -13.0 (-13.1, -12.9) | -69.0 (-69.1, -68.9) | -5.0 (-5.14, -4.86) | 14.0 (13.86, 14.14) | -44.0 (-44.12, -43.88) | -25.0 (-25.11, -24.89) |
| vicuna-13b-v1.5 | 2.0 (1.93, 2.07) | -4.0 (-4.14, -3.86) | 10.0 (9.91, 10.09) | -3.0 (-3.08, -2.92) | -3.0 (-3.08, -2.92) | 15.0 (14.88, 15.12) | 33.0 (32.87, 33.13) | -2.0 (-2.11, -1.89) | -5.0 (-5.08, -4.92) |
| vicuna-13b-v1.5-w4-g128 | 0.0 (-0.08, 0.08) | -5.0 (-5.14, -4.86) | 11.0 (10.91, 11.09) | -4.0 (-4.08, -3.92) | 0.0 (-0.07, 0.07) | 6.0 (5.87, 6.13) | 31.0 (30.87, 31.13) | 2.0 (1.89, 2.11) | -4.0 (-4.08, -3.92) |
| WizardLM-13B-V1.2 | -3.0 (-3.08, -2.92) | 17.0 (16.86, 17.14) | -4.0 (-4.11, -3.89) | -16.0 (-16.1, -15.9) | -14.0 (-14.1, -13.9) | -7.0 (-7.14, -6.86) | 13.0 (12.86, 13.14) | -16.0 (-16.12, -15.88) | -15.0 (-15.1, -14.9) |
| WizardLM-13B-V1.2-w4-g128 | -2.0 (-2.08, -1.92) | 16.0 (15.86, 16.14) | 8.0 (7.91, 8.09) | -2.0 (-2.08, -1.92) | 0.0 (-0.07, 0.07) | 5.0 (4.87, 5.13) | 24.0 (23.86, 24.14) | -11.0 (-11.12, -10.88) | -5.0 (-5.08, -4.92) |
| Llama-2-13b-chat-hf | -65.0 (-65.1, -64.9) | -34.0 (-34.13, -33.87) | -51.0 (-51.12, -50.88) | -62.0 (-62.11, -61.89) | -54.0 (-54.11, -53.89) | -35.0 (-35.13, -34.87) | -18.0 (-18.13, -17.87) | -43.0 (-43.12, -42.88) | -37.0 (-37.11, -36.89) |
| Llama-2-13b-chat-hf-w4-g128 | -41.0 (-41.11, -40.89) | -39.0 (-39.12, -38.88) | -16.0 (-16.12, -15.88) | -36.0 (-36.11, -35.89) | -42.0 (-42.11, -41.89) | -29.0 (-29.14, -28.86) | -5.0 (-5.14, -4.86) | -41.0 (-41.12, -40.88) | -21.0 (-21.1, -20.9) |
| vicuna-33b-v1.3 | -31.0 (-31.11, -30.89) | 5.0 (4.86, 5.14) | 7.0 (6.9, 7.1) | 1.0 (0.93, 1.07) | -10.0 (-10.09, -9.91) | 15.0 (14.88, 15.12) | 24.0 (23.86, 24.14) | -7.0 (-7.12, -6.88) | 1.0 (0.93, 1.07) |
| vicuna-33b-v1.3-w4-g128 | -33.0 (-33.11, -32.89) | 13.0 (12.86, 13.14) | 5.0 (4.9, 5.1) | 1.0 (0.93, 1.07) | -9.0 (-9.09, -8.91) | 3.0 (2.87, 3.13) | 23.0 (22.86, 23.14) | -5.0 (-5.11, -4.89) | -1.0 (-1.07, -0.93) |

**Table S8 (a). Model Accuracy_mentioned_ on each SDoH question (Refined pipeline)**

| **Model** | **Q1** | **Q2** | **Q3** | **Q4** | **Q5** | **Q6** | **Q7** | **Q8** | **Q9** |
| --- | --- | --- | --- | --- | --- | --- | --- | --- | --- |
| openchat_3.5 | 95.6 (95.56, 95.65) | 78.33 (78.15, 78.52) | 100.0 (100.0, 100.0) | 69.23 (66.81, 71.65) | 87.5 (83.28, 91.72) | 77.05 (76.87, 77.23) | 73.53 (73.37, 73.69) | 78.26 (77.43, 79.09) | 90.0 (87.49, 92.51) |
| openchat_3.5-w4-g128 | 96.7 (96.66, 96.74) | 73.33 (73.14, 73.53) | 100.0 (100.0, 100.0) | 76.92 (74.71, 79.13) | 75.0 (69.47, 80.53) | 81.97 (81.8, 82.13) | 75.0 (74.84, 75.16) | 73.91 (73.03, 74.8) | 90.0 (87.49, 92.51) |
| zephyr-7b-beta | 69.23 (69.12, 69.34) | 35.0 (34.79, 35.21) | 29.41 (27.9, 30.92) | 69.23 (66.81, 71.65) | 12.5 (8.28, 16.72) | 24.59 (24.4, 24.78) | 51.47 (51.29, 51.65) | 30.43 (29.51, 31.36) | 30.0 (26.16, 33.84) |
| zephyr-7b-beta-w4-g128 | 72.53 (72.42, 72.63) | 25.0 (24.81, 25.19) | 23.53 (22.12, 24.93) | 38.46 (35.91, 41.01) | 25.0 (19.47, 30.53) | 22.95 (22.77, 23.13) | 41.18 (41.0, 41.36) | 21.74 (20.91, 22.57) | 40.0 (35.9, 44.1) |
| vicuna-7b-v1.5 | 81.32 (81.23, 81.41) | 65.0 (64.79, 65.21) | 23.53 (22.12, 24.93) | 23.08 (20.87, 25.29) | 25.0 (19.47, 30.53) | 4.92 (4.82, 5.01) | 47.06 (46.88, 47.24) | 56.52 (55.53, 57.52) | 20.0 (16.65, 23.35) |
| vicuna-7b-v1.5-w4-g128 | 76.92 (76.83, 77.02) | 63.33 (63.12, 63.55) | 23.53 (22.12, 24.93) | 7.69 (6.3, 9.09) | 12.5 (8.28, 16.72) | 4.92 (4.82, 5.01) | 52.94 (52.76, 53.12) | 39.13 (38.15, 40.11) | 30.0 (26.16, 33.84) |
| Llama-2-7b-chat-hf | 62.64 (62.52, 62.75) | 41.67 (41.45, 41.88) | 64.71 (63.12, 66.29) | 15.38 (13.49, 17.28) | 50.0 (43.62, 56.38) | 14.75 (14.6, 14.91) | 30.88 (30.71, 31.05) | 52.17 (51.17, 53.18) | 70.0 (66.16, 73.84) |
| Llama-2-7b-chat-hf-w4-g128 | 90.11 (90.04, 90.18) | 28.33 (28.13, 28.53) | 82.35 (81.09, 83.62) | 15.38 (13.49, 17.28) | 87.5 (83.28, 91.72) | 13.11 (12.97, 13.26) | 39.71 (39.53, 39.88) | 56.52 (55.53, 57.52) | 60.0 (55.9, 64.1) |
| vicuna-13b-v1.5 | 94.51 (94.45, 94.56) | 70.0 (69.8, 70.2) | 76.47 (75.07, 77.88) | 23.08 (20.87, 25.29) | 12.5 (8.28, 16.72) | 39.34 (39.13, 39.55) | 38.24 (38.06, 38.41) | 56.52 (55.53, 57.52) | 90.0 (87.49, 92.51) |
| vicuna-13b-v1.5-w4-g128 | 91.21 (91.14, 91.27) | 68.33 (68.13, 68.54) | 94.12 (93.34, 94.9) | 23.08 (20.87, 25.29) | 37.5 (31.32, 43.68) | 26.23 (26.04, 26.42) | 35.29 (35.12, 35.47) | 65.22 (64.26, 66.17) | 80.0 (76.65, 83.35) |
| WizardLM-13B-V1.2 | 92.31 (92.25, 92.37) | 70.0 (69.8, 70.2) | 58.82 (57.19, 60.45) | 46.15 (43.54, 48.77) | 25.0 (19.47, 30.53) | 22.95 (22.77, 23.13) | 52.94 (52.76, 53.12) | 65.22 (64.26, 66.17) | 90.0 (87.49, 92.51) |
| WizardLM-13B-V1.2-w4-g128 | 91.21 (91.14, 91.27) | 70.0 (69.8, 70.2) | 58.82 (57.19, 60.45) | 30.77 (28.35, 33.19) | 25.0 (19.47, 30.53) | 40.98 (40.77, 41.2) | 51.47 (51.29, 51.65) | 65.22 (64.26, 66.17) | 90.0 (87.49, 92.51) |
| Llama-2-13b-chat-hf | 36.26 (36.15, 36.38) | 10.0 (9.87, 10.13) | 11.76 (10.7, 12.83) | 7.69 (6.3, 9.09) | 12.5 (8.28, 16.72) | 14.75 (14.6, 14.91) | 22.06 (21.91, 22.21) | 30.43 (29.51, 31.36) | 30.0 (26.16, 33.84) |
| Llama-2-13b-chat-hf-w4-g128 | 53.85 (53.73, 53.96) | 6.67 (6.56, 6.78) | 29.41 (27.9, 30.92) | 7.69 (6.3, 9.09) | 12.5 (8.28, 16.72) | 11.48 (11.34, 11.61) | 10.29 (10.18, 10.4) | 47.83 (46.82, 48.83) | 60.0 (55.9, 64.1) |
| vicuna-33b-v1.3 | 57.14 (57.03, 57.26) | 68.33 (68.13, 68.54) | 52.94 (51.29, 54.59) | 53.85 (51.23, 56.46) | 75.0 (69.47, 80.53) | 4.92 (4.82, 5.01) | 66.18 (66.0, 66.35) | 56.52 (55.53, 57.52) | 50.0 (45.81, 54.19) |
| vicuna-33b-v1.3-w4-g128 | 54.95 (54.83, 55.06) | 70.0 (69.8, 70.2) | 41.18 (39.55, 42.81) | 53.85 (51.23, 56.46) | 37.5 (31.32, 43.68) | 3.28 (3.2, 3.36) | 63.24 (63.06, 63.41) | 52.17 (51.17, 53.18) | 40.0 (35.9, 44.1) |
| baseline | 91.21 (91.14, 91.27) | 18.33 (18.16, 18.5) | 0.0 (0.0, 0.0) | 38.46 (35.91, 41.01) | 12.5 (8.28, 16.72) | 49.18 (48.97, 49.4) | 20.59 (20.44, 20.74) | 60.87 (59.89, 61.85) | 60.0 (55.9, 64.1) |

**Table S8 (b). Model Accuracy_mentioned_ on each SDoH question (Default pipeline)**

| **Model** | **Q1** | **Q2** | **Q3** | **Q4** | **Q5** | **Q6** | **Q7** | **Q8** | **Q9** |
| --- | --- | --- | --- | --- | --- | --- | --- | --- | --- |
| openchat_3.5 | 95.6 (95.56, 95.65) | 78.33 (78.15, 78.52) | 100.0 (100.0, 100.0) | 69.23 (66.81, 71.65) | 87.5 (83.28, 91.72) | 91.8 (91.69, 91.92) | 61.76 (61.59, 61.94) | 73.91 (73.03, 74.8) | 90.0 (87.49, 92.51) |
| openchat_3.5-w4-g128 | 96.7 (96.66, 96.74) | 76.67 (76.48, 76.85) | 100.0 (100.0, 100.0) | 76.92 (74.71, 79.13) | 75.0 (69.47, 80.53) | 93.44 (93.34, 93.55) | 58.82 (58.64, 59.0) | 73.91 (73.03, 74.8) | 90.0 (87.49, 92.51) |
| zephyr-7b-beta | 69.23 (69.12, 69.34) | 41.67 (41.45, 41.88) | 29.41 (27.9, 30.92) | 69.23 (66.81, 71.65) | 12.5 (8.28, 16.72) | 49.18 (48.97, 49.4) | 38.24 (38.06, 38.41) | 30.43 (29.51, 31.36) | 30.0 (26.16, 33.84) |
| zephyr-7b-beta-w4-g128 | 72.53 (72.42, 72.63) | 26.67 (26.47, 26.86) | 23.53 (22.12, 24.93) | 38.46 (35.91, 41.01) | 25.0 (19.47, 30.53) | 24.59 (24.4, 24.78) | 47.06 (46.88, 47.24) | 21.74 (20.91, 22.57) | 30.0 (26.16, 33.84) |
| vicuna-7b-v1.5 | 80.22 (80.13, 80.31) | 78.33 (78.15, 78.52) | 23.53 (22.12, 24.93) | 23.08 (20.87, 25.29) | 25.0 (19.47, 30.53) | 44.26 (44.05, 44.48) | 25.0 (24.84, 25.16) | 56.52 (55.53, 57.52) | 20.0 (16.65, 23.35) |
| vicuna-7b-v1.5-w4-g128 | 75.82 (75.72, 75.92) | 75.0 (74.81, 75.19) | 23.53 (22.12, 24.93) | 7.69 (6.3, 9.09) | 12.5 (8.28, 16.72) | 36.07 (35.86, 36.27) | 20.59 (20.44, 20.74) | 39.13 (38.15, 40.11) | 30.0 (26.16, 33.84) |
| Llama-2-7b-chat-hf | 63.74 (63.62, 63.85) | 46.67 (46.45, 46.89) | 64.71 (63.12, 66.29) | 7.69 (6.3, 9.09) | 62.5 (56.32, 68.68) | 49.18 (48.97, 49.4) | 35.29 (35.12, 35.47) | 65.22 (64.26, 66.17) | 50.0 (45.81, 54.19) |
| Llama-2-7b-chat-hf-w4-g128 | 87.91 (87.84, 87.99) | 66.67 (66.46, 66.87) | 82.35 (81.09, 83.62) | 15.38 (13.49, 17.28) | 87.5 (83.28, 91.72) | 70.49 (70.3, 70.69) | 52.94 (52.76, 53.12) | 56.52 (55.53, 57.52) | 60.0 (55.9, 64.1) |
| vicuna-13b-v1.5 | 93.41 (93.35, 93.46) | 78.33 (78.15, 78.52) | 76.47 (75.07, 77.88) | 23.08 (20.87, 25.29) | 12.5 (8.28, 16.72) | 91.8 (91.69, 91.92) | 66.18 (66.0, 66.35) | 47.83 (46.82, 48.83) | 90.0 (87.49, 92.51) |
| vicuna-13b-v1.5-w4-g128 | 91.21 (91.14, 91.27) | 75.0 (74.81, 75.19) | 88.24 (87.17, 89.3) | 23.08 (20.87, 25.29) | 37.5 (31.32, 43.68) | 81.97 (81.8, 82.13) | 64.71 (64.53, 64.88) | 56.52 (55.53, 57.52) | 80.0 (76.65, 83.35) |
| WizardLM-13B-V1.2 | 92.31 (92.25, 92.37) | 78.33 (78.15, 78.52) | 58.82 (57.19, 60.45) | 46.15 (43.54, 48.77) | 25.0 (19.47, 30.53) | 83.61 (83.45, 83.77) | 72.06 (71.9, 72.22) | 60.87 (59.89, 61.85) | 90.0 (87.49, 92.51) |
| WizardLM-13B-V1.2-w4-g128 | 91.21 (91.14, 91.27) | 76.67 (76.48, 76.85) | 58.82 (57.19, 60.45) | 30.77 (28.35, 33.19) | 25.0 (19.47, 30.53) | 90.16 (90.04, 90.29) | 73.53 (73.37, 73.69) | 56.52 (55.53, 57.52) | 90.0 (87.49, 92.51) |
| Llama-2-13b-chat-hf | 26.37 (26.27, 26.48) | 21.67 (21.48, 21.85) | 11.76 (10.7, 12.83) | 7.69 (6.3, 9.09) | 0.0 (0.0, 0.0) | 16.39 (16.23, 16.55) | 25.0 (24.84, 25.16) | 26.09 (25.2, 26.97) | 60.0 (55.9, 64.1) |
| Llama-2-13b-chat-hf-w4-g128 | 52.75 (52.63, 52.86) | 8.33 (8.21, 8.46) | 29.41 (27.9, 30.92) | 7.69 (6.3, 9.09) | 0.0 (0.0, 0.0) | 9.84 (9.71, 9.96) | 29.41 (29.25, 29.58) | 47.83 (46.82, 48.83) | 60.0 (55.9, 64.1) |
| vicuna-33b-v1.3 | 57.14 (57.03, 57.26) | 76.67 (76.48, 76.85) | 52.94 (51.29, 54.59) | 53.85 (51.23, 56.46) | 75.0 (69.47, 80.53) | 73.77 (73.58, 73.96) | 57.35 (57.17, 57.53) | 56.52 (55.53, 57.52) | 50.0 (45.81, 54.19) |
| vicuna-33b-v1.3-w4-g128 | 54.95 (54.83, 55.06) | 76.67 (76.48, 76.85) | 41.18 (39.55, 42.81) | 53.85 (51.23, 56.46) | 37.5 (31.32, 43.68) | 49.18 (48.97, 49.4) | 55.88 (55.7, 56.06) | 47.83 (46.82, 48.83) | 40.0 (35.9, 44.1) |
| baseline | 91.21 (91.14, 91.27) | 18.33 (18.16, 18.5) | 0.0 (0.0, 0.0) | 38.46 (35.91, 41.01) | 12.5 (8.28, 16.72) | 44.26 (44.05, 44.48) | 20.59 (20.44, 20.74) | 60.87 (59.89, 61.85) | 60.0 (55.9, 64.1) |

**Table S9 (a). Model Accuracy_mentioned_ difference (compared to the baseline) on each SDoH question (Refined pipeline)**

| **Model** | **Q1** | **Q2** | **Q3** | **Q4** | **Q5** | **Q6** | **Q7** | **Q8** | **Q9** |
| --- | --- | --- | --- | --- | --- | --- | --- | --- | --- |
| openchat_3.5 | 4.4 (4.31, 4.48) | 60.0 (59.75, 60.25) | 100.0 (100.0, 100.0) | 30.77 (27.44, 34.1) | 75.0 (69.58, 80.42) | 27.87 (27.59, 28.15) | 52.94 (52.73, 53.16) | 17.39 (16.14, 18.64) | 30.0 (25.46, 34.54) |
| openchat_3.5-w4-g128 | 5.49 (5.42, 5.57) | 55.0 (54.74, 55.26) | 100.0 (100.0, 100.0) | 38.46 (35.26, 41.66) | 62.5 (56.15, 68.85) | 32.79 (32.52, 33.06) | 54.41 (54.2, 54.63) | 13.04 (11.76, 14.33) | 30.0 (25.46, 34.54) |
| zephyr-7b-beta | -21.98 (-22.1, -21.85) | 16.67 (16.4, 16.94) | 29.41 (27.9, 30.92) | 30.77 (27.44, 34.1) | 0.0 (-5.42, 5.42) | -24.59 (-24.87, -24.31) | 30.88 (30.65, 31.11) | -30.43 (-31.74, -29.12) | -30.0 (-35.22, -24.78) |
| zephyr-7b-beta-w4-g128 | -18.68 (-18.8, -18.56) | 6.67 (6.41, 6.92) | 23.53 (22.12, 24.93) | 0.0 (-3.42, 3.42) | 12.5 (6.15, 18.85) | -26.23 (-26.51, -25.95) | 20.59 (20.36, 20.82) | -39.13 (-40.38, -37.88) | -20.0 (-25.39, -14.61) |
| vicuna-7b-v1.5 | -9.89 (-10.0, -9.78) | 46.67 (46.4, 46.94) | 23.53 (22.12, 24.93) | -15.38 (-18.58, -12.19) | 12.5 (6.15, 18.85) | -44.26 (-44.5, -44.03) | 26.47 (26.24, 26.7) | -4.35 (-5.71, -2.99) | -40.0 (-44.94, -35.06) |
| vicuna-7b-v1.5-w4-g128 | -14.29 (-14.4, -14.17) | 45.0 (44.73, 45.27) | 23.53 (22.12, 24.93) | -30.77 (-33.57, -27.97) | 0.0 (-5.42, 5.42) | -44.26 (-44.5, -44.03) | 32.35 (32.12, 32.58) | -21.74 (-23.09, -20.39) | -30.0 (-35.22, -24.78) |
| Llama-2-7b-chat-hf | -28.57 (-28.7, -28.44) | 23.33 (23.06, 23.61) | 64.71 (63.12, 66.29) | -23.08 (-26.1, -20.06) | 37.5 (30.46, 44.54) | -34.43 (-34.69, -34.16) | 10.29 (10.07, 10.52) | -8.7 (-10.06, -7.33) | 10.0 (4.78, 15.22) |
| Llama-2-7b-chat-hf-w4-g128 | -1.1 (-1.19, -1.0) | 10.0 (9.74, 10.26) | 82.35 (81.09, 83.62) | -23.08 (-26.1, -20.06) | 75.0 (69.58, 80.42) | -36.07 (-36.32, -35.81) | 19.12 (18.89, 19.35) | -4.35 (-5.71, -2.99) | 0.0 (-5.39, 5.39) |
| vicuna-13b-v1.5 | 3.3 (3.21, 3.38) | 51.67 (51.4, 51.93) | 76.47 (75.07, 77.88) | -15.38 (-18.58, -12.19) | 0.0 (-5.42, 5.42) | -9.84 (-10.13, -9.54) | 17.65 (17.42, 17.88) | -4.35 (-5.71, -2.99) | 30.0 (25.46, 34.54) |
| vicuna-13b-v1.5-w4-g128 | 0.0 (-0.09, 0.09) | 50.0 (49.74, 50.26) | 94.12 (93.34, 94.9) | -15.38 (-18.58, -12.19) | 25.0 (18.12, 31.88) | -22.95 (-23.23, -22.67) | 14.71 (14.48, 14.93) | 4.35 (3.02, 5.68) | 20.0 (15.06, 24.94) |
| WizardLM-13B-V1.2 | 1.1 (1.01, 1.19) | 51.67 (51.4, 51.93) | 58.82 (57.19, 60.45) | 7.69 (4.23, 11.15) | 12.5 (6.15, 18.85) | -26.23 (-26.51, -25.95) | 32.35 (32.12, 32.58) | 4.35 (3.02, 5.68) | 30.0 (25.46, 34.54) |
| WizardLM-13B-V1.2-w4-g128 | 0.0 (-0.09, 0.09) | 51.67 (51.4, 51.93) | 58.82 (57.19, 60.45) | -7.69 (-11.02, -4.36) | 12.5 (6.15, 18.85) | -8.2 (-8.5, -7.9) | 30.88 (30.65, 31.11) | 4.35 (3.02, 5.68) | 30.0 (25.46, 34.54) |
| Llama-2-13b-chat-hf | -54.95 (-55.07, -54.82) | -8.33 (-8.55, -8.12) | 11.76 (10.7, 12.83) | -30.77 (-33.57, -27.97) | 0.0 (-5.42, 5.42) | -34.43 (-34.69, -34.16) | 1.47 (1.26, 1.68) | -30.43 (-31.74, -29.12) | -30.0 (-35.22, -24.78) |
| Llama-2-13b-chat-hf-w4-g128 | -37.36 (-37.5, -37.23) | -11.67 (-11.87, -11.47) | 29.41 (27.9, 30.92) | -30.77 (-33.57, -27.97) | 0.0 (-5.42, 5.42) | -37.7 (-37.96, -37.45) | -10.29 (-10.48, -10.11) | -13.04 (-14.41, -11.68) | 0.0 (-5.39, 5.39) |
| vicuna-33b-v1.3 | -34.07 (-34.2, -33.93) | 50.0 (49.74, 50.26) | 52.94 (51.29, 54.59) | 15.38 (11.93, 18.84) | 62.5 (56.15, 68.85) | -44.26 (-44.5, -44.03) | 45.59 (45.36, 45.81) | -4.35 (-5.71, -2.99) | -10.0 (-15.45, -4.55) |
| vicuna-33b-v1.3-w4-g128 | -36.26 (-36.4, -36.13) | 51.67 (51.4, 51.93) | 41.18 (39.55, 42.81) | 15.38 (11.93, 18.84) | 25.0 (18.12, 31.88) | -45.9 (-46.13, -45.67) | 42.65 (42.42, 42.87) | -8.7 (-10.06, -7.33) | -20.0 (-25.39, -14.61) |

**Table S9 (b). Model Accuracy_mentioned_ difference (compared to the baseline) on each SDoH question (Default pipeline)**

| **Model** | **Q1** | **Q2** | **Q3** | **Q4** | **Q5** | **Q6** | **Q7** | **Q8** | **Q9** |
| --- | --- | --- | --- | --- | --- | --- | --- | --- | --- |
| openchat_3.5 | 4.4 (4.31, 4.48) | 60.0 (59.75, 60.25) | 100.0 (100.0, 100.0) | 30.77 (27.44, 34.1) | 75.0 (69.58, 80.42) | 47.54 (47.3, 47.78) | 41.18 (40.95, 41.4) | 13.04 (11.76, 14.33) | 30.0 (25.46, 34.54) |
| openchat_3.5-w4-g128 | 5.49 (5.42, 5.57) | 58.33 (58.08, 58.58) | 100.0 (100.0, 100.0) | 38.46 (35.26, 41.66) | 62.5 (56.15, 68.85) | 49.18 (48.94, 49.42) | 38.24 (38.01, 38.47) | 13.04 (11.76, 14.33) | 30.0 (25.46, 34.54) |
| zephyr-7b-beta | -21.98 (-22.1, -21.85) | 23.33 (23.06, 23.61) | 29.41 (27.9, 30.92) | 30.77 (27.44, 34.1) | 0.0 (-5.42, 5.42) | 4.92 (4.62, 5.22) | 17.65 (17.42, 17.88) | -30.43 (-31.74, -29.12) | -30.0 (-35.22, -24.78) |
| zephyr-7b-beta-w4-g128 | -18.68 (-18.8, -18.56) | 8.33 (8.08, 8.59) | 23.53 (22.12, 24.93) | 0.0 (-3.42, 3.42) | 12.5 (6.15, 18.85) | -19.67 (-19.95, -19.39) | 26.47 (26.24, 26.7) | -39.13 (-40.38, -37.88) | -30.0 (-35.22, -24.78) |
| vicuna-7b-v1.5 | -10.99 (-11.1, -10.88) | 60.0 (59.75, 60.25) | 23.53 (22.12, 24.93) | -15.38 (-18.58, -12.19) | 12.5 (6.15, 18.85) | 0.0 (-0.3, 0.3) | 4.41 (4.2, 4.63) | -4.35 (-5.71, -2.99) | -40.0 (-44.94, -35.06) |
| vicuna-7b-v1.5-w4-g128 | -15.38 (-15.5, -15.27) | 56.67 (56.41, 56.92) | 23.53 (22.12, 24.93) | -30.77 (-33.57, -27.97) | 0.0 (-5.42, 5.42) | -8.2 (-8.49, -7.9) | 0.0 (-0.21, 0.21) | -21.74 (-23.09, -20.39) | -30.0 (-35.22, -24.78) |
| Llama-2-7b-chat-hf | -27.47 (-27.6, -27.34) | 28.33 (28.06, 28.61) | 64.71 (63.12, 66.29) | -30.77 (-33.57, -27.97) | 50.0 (43.12, 56.88) | 4.92 (4.62, 5.22) | 14.71 (14.48, 14.93) | 4.35 (3.02, 5.68) | -10.0 (-15.45, -4.55) |
| Llama-2-7b-chat-hf-w4-g128 | -3.3 (-3.4, -3.2) | 48.33 (48.07, 48.6) | 82.35 (81.09, 83.62) | -23.08 (-26.1, -20.06) | 75.0 (69.58, 80.42) | 26.23 (25.94, 26.52) | 32.35 (32.12, 32.58) | -4.35 (-5.71, -2.99) | 0.0 (-5.39, 5.39) |
| vicuna-13b-v1.5 | 2.2 (2.11, 2.28) | 60.0 (59.75, 60.25) | 76.47 (75.07, 77.88) | -15.38 (-18.58, -12.19) | 0.0 (-5.42, 5.42) | 47.54 (47.3, 47.78) | 45.59 (45.36, 45.81) | -13.04 (-14.41, -11.68) | 30.0 (25.46, 34.54) |
| vicuna-13b-v1.5-w4-g128 | 0.0 (-0.09, 0.09) | 56.67 (56.41, 56.92) | 88.24 (87.17, 89.3) | -15.38 (-18.58, -12.19) | 25.0 (18.12, 31.88) | 37.7 (37.44, 37.97) | 44.12 (43.89, 44.34) | -4.35 (-5.71, -2.99) | 20.0 (15.06, 24.94) |
| WizardLM-13B-V1.2 | 1.1 (1.01, 1.19) | 60.0 (59.75, 60.25) | 58.82 (57.19, 60.45) | 7.69 (4.23, 11.15) | 12.5 (6.15, 18.85) | 39.34 (39.08, 39.61) | 51.47 (51.25, 51.69) | 0.0 (-1.35, 1.35) | 30.0 (25.46, 34.54) |
| WizardLM-13B-V1.2-w4-g128 | 0.0 (-0.09, 0.09) | 58.33 (58.08, 58.58) | 58.82 (57.19, 60.45) | -7.69 (-11.02, -4.36) | 12.5 (6.15, 18.85) | 45.9 (45.65, 46.15) | 52.94 (52.73, 53.16) | -4.35 (-5.71, -2.99) | 30.0 (25.46, 34.54) |
| Llama-2-13b-chat-hf | -64.84 (-64.96, -64.71) | 3.33 (3.09, 3.58) | 11.76 (10.7, 12.83) | -30.77 (-33.57, -27.97) | -12.5 (-16.72, -8.28) | -27.87 (-28.13, -27.6) | 4.41 (4.2, 4.63) | -34.78 (-36.07, -33.5) | 0.0 (-5.39, 5.39) |
| Llama-2-13b-chat-hf-w4-g128 | -38.46 (-38.59, -38.33) | -10.0 (-10.21, -9.79) | 29.41 (27.9, 30.92) | -30.77 (-33.57, -27.97) | -12.5 (-16.72, -8.28) | -34.43 (-34.67, -34.18) | 8.82 (8.6, 9.04) | -13.04 (-14.41, -11.68) | 0.0 (-5.39, 5.39) |
| vicuna-33b-v1.3 | -34.07 (-34.2, -33.93) | 58.33 (58.08, 58.58) | 52.94 (51.29, 54.59) | 15.38 (11.93, 18.84) | 62.5 (56.15, 68.85) | 29.51 (29.23, 29.79) | 36.76 (36.53, 37.0) | -4.35 (-5.71, -2.99) | -10.0 (-15.45, -4.55) |
| vicuna-33b-v1.3-w4-g128 | -36.26 (-36.4, -36.13) | 58.33 (58.08, 58.58) | 41.18 (39.55, 42.81) | 15.38 (11.93, 18.84) | 25.0 (18.12, 31.88) | 4.92 (4.62, 5.22) | 35.29 (35.06, 35.53) | -13.04 (-14.41, -11.68) | -20.0 (-25.39, -14.61) |

**Table S10 (a). Model Accuracy_non-mentioned_ on each SDoH question (Refined pipeline)**

| **Model** | **Q1** | **Q2** | **Q3** | **Q4** | **Q5** | **Q6** | **Q7** | **Q8** | **Q9** |
| --- | --- | --- | --- | --- | --- | --- | --- | --- | --- |
| openchat_3.5 | 100.0 (100.0, 100.0) | 90.0 (89.75, 90.25) | 95.18 (95.12, 95.24) | 96.55 (96.51, 96.6) | 97.83 (97.79, 97.86) | 97.44 (97.3, 97.57) | 87.5 (87.11, 87.89) | 98.7 (98.67, 98.74) | 97.78 (97.74, 97.81) |
| openchat_3.5-w4-g128 | 88.89 (85.69, 92.09) | 90.0 (89.75, 90.25) | 95.18 (95.12, 95.24) | 97.7 (97.66, 97.74) | 97.83 (97.79, 97.86) | 92.31 (92.08, 92.54) | 87.5 (87.11, 87.89) | 97.4 (97.35, 97.45) | 97.78 (97.74, 97.81) |
| zephyr-7b-beta | 100.0 (100.0, 100.0) | 97.5 (97.37, 97.63) | 92.77 (92.7, 92.84) | 89.66 (89.58, 89.73) | 84.78 (84.7, 84.86) | 69.23 (68.83, 69.63) | 84.38 (83.95, 84.8) | 81.82 (81.7, 81.93) | 94.44 (94.39, 94.5) |
| zephyr-7b-beta-w4-g128 | 100.0 (100.0, 100.0) | 100.0 (100.0, 100.0) | 68.67 (68.55, 68.8) | 65.52 (65.4, 65.64) | 72.83 (72.72, 72.93) | 84.62 (84.3, 84.93) | 68.75 (68.2, 69.3) | 74.03 (73.89, 74.16) | 95.56 (95.51, 95.6) |
| vicuna-7b-v1.5 | 100.0 (100.0, 100.0) | 77.5 (77.15, 77.85) | 98.8 (98.77, 98.82) | 98.85 (98.82, 98.88) | 97.83 (97.79, 97.86) | 100.0 (100.0, 100.0) | 96.88 (96.67, 97.08) | 66.23 (66.09, 66.38) | 64.44 (64.33, 64.56) |
| vicuna-7b-v1.5-w4-g128 | 100.0 (100.0, 100.0) | 62.5 (62.1, 62.9) | 97.59 (97.55, 97.63) | 98.85 (98.82, 98.88) | 100.0 (100.0, 100.0) | 94.87 (94.68, 95.06) | 90.62 (90.28, 90.97) | 92.21 (92.13, 92.29) | 84.44 (84.36, 84.53) |
| Llama-2-7b-chat-hf | 33.33 (28.53, 38.14) | 82.5 (82.18, 82.82) | 74.7 (74.58, 74.82) | 88.51 (88.43, 88.59) | 47.83 (47.71, 47.94) | 69.23 (68.83, 69.63) | 6.25 (5.96, 6.54) | 29.87 (29.73, 30.01) | 83.33 (83.25, 83.42) |
| Llama-2-7b-chat-hf-w4-g128 | 44.44 (39.38, 49.51) | 95.0 (94.82, 95.18) | 59.04 (58.9, 59.17) | 89.66 (89.58, 89.73) | 20.65 (20.56, 20.74) | 69.23 (68.83, 69.63) | 12.5 (12.11, 12.89) | 35.06 (34.92, 35.21) | 68.89 (68.78, 69.0) |
| vicuna-13b-v1.5 | 100.0 (100.0, 100.0) | 90.0 (89.75, 90.25) | 97.59 (97.55, 97.63) | 100.0 (100.0, 100.0) | 97.83 (97.79, 97.86) | 100.0 (100.0, 100.0) | 21.88 (21.39, 22.36) | 92.21 (92.13, 92.29) | 88.89 (88.81, 88.96) |
| vicuna-13b-v1.5-w4-g128 | 100.0 (100.0, 100.0) | 80.0 (79.67, 80.33) | 96.39 (96.34, 96.44) | 98.85 (98.82, 98.88) | 98.91 (98.89, 98.94) | 100.0 (100.0, 100.0) | 59.38 (58.79, 59.96) | 94.81 (94.74, 94.87) | 91.11 (91.04, 91.18) |
| WizardLM-13B-V1.2 | 55.56 (50.49, 60.62) | 87.5 (87.23, 87.77) | 83.13 (83.03, 83.23) | 81.61 (81.51, 81.71) | 84.78 (84.7, 84.86) | 94.87 (94.68, 95.06) | 84.38 (83.95, 84.8) | 75.32 (75.2, 75.45) | 77.78 (77.68, 77.88) |
| WizardLM-13B-V1.2-w4-g128 | 77.78 (73.54, 82.01) | 85.0 (84.7, 85.3) | 97.59 (97.55, 97.63) | 98.85 (98.82, 98.88) | 98.91 (98.89, 98.94) | 94.87 (94.68, 95.06) | 65.62 (65.06, 66.19) | 76.62 (76.5, 76.75) | 87.78 (87.7, 87.86) |
| Llama-2-13b-chat-hf | 33.33 (28.53, 38.14) | 67.5 (67.11, 67.89) | 51.81 (51.67, 51.94) | 29.89 (29.77, 30.0) | 50.0 (49.89, 50.11) | 43.59 (43.16, 44.02) | 28.12 (27.59, 28.66) | 42.86 (42.71, 43.01) | 72.22 (72.12, 72.33) |
| Llama-2-13b-chat-hf-w4-g128 | 44.44 (39.38, 49.51) | 65.0 (64.6, 65.4) | 75.9 (75.79, 76.02) | 63.22 (63.1, 63.34) | 69.57 (69.46, 69.67) | 51.28 (50.85, 51.71) | 28.12 (27.59, 28.66) | 51.95 (51.8, 52.1) | 88.89 (88.81, 88.96) |
| vicuna-33b-v1.3 | 100.0 (100.0, 100.0) | 92.5 (92.28, 92.72) | 97.59 (97.55, 97.63) | 100.0 (100.0, 100.0) | 83.7 (83.61, 83.78) | 100.0 (100.0, 100.0) | 34.38 (33.81, 34.94) | 83.12 (83.0, 83.23) | 98.89 (98.86, 98.91) |
| vicuna-33b-v1.3-w4-g128 | 100.0 (100.0, 100.0) | 92.5 (92.28, 92.72) | 97.59 (97.55, 97.63) | 98.85 (98.82, 98.88) | 89.13 (89.06, 89.2) | 100.0 (100.0, 100.0) | 28.12 (27.59, 28.66) | 87.01 (86.91, 87.11) | 98.89 (98.86, 98.91) |
| baseline | 100.0 (100.0, 100.0) | 100.0 (100.0, 100.0) | 100.0 (100.0, 100.0) | 100.0 (100.0, 100.0) | 100.0 (100.0, 100.0) | 97.44 (97.3, 97.57) | 87.5 (87.11, 87.89) | 88.31 (88.22, 88.41) | 96.67 (96.62, 96.71) |

**Table S10 (b). Model Accuracy_non-mentioned_ on each SDoH question (Default pipeline)**

| **Model** | **Q1** | **Q2** | **Q3** | **Q4** | **Q5** | **Q6** | **Q7** | **Q8** | **Q9** |
| --- | --- | --- | --- | --- | --- | --- | --- | --- | --- |
| openchat_3.5 | 100.0 (100.0, 100.0) | 62.5 (62.1, 62.9) | 95.18 (95.12, 95.24) | 96.55 (96.51, 96.6) | 96.74 (96.7, 96.78) | 51.28 (50.85, 51.71) | 100.0 (100.0, 100.0) | 97.4 (97.35, 97.45) | 97.78 (97.74, 97.81) |
| openchat_3.5-w4-g128 | 88.89 (85.69, 92.09) | 57.5 (57.09, 57.91) | 95.18 (95.12, 95.24) | 97.7 (97.66, 97.74) | 96.74 (96.7, 96.78) | 46.15 (45.72, 46.58) | 100.0 (100.0, 100.0) | 96.1 (96.05, 96.16) | 97.78 (97.74, 97.81) |
| zephyr-7b-beta | 88.89 (85.69, 92.09) | 87.5 (87.23, 87.77) | 89.16 (89.07, 89.24) | 86.21 (86.12, 86.29) | 79.35 (79.26, 79.44) | 87.18 (86.89, 87.47) | 96.88 (96.67, 97.08) | 63.64 (63.49, 63.78) | 92.22 (92.16, 92.29) |
| zephyr-7b-beta-w4-g128 | 100.0 (100.0, 100.0) | 87.5 (87.23, 87.77) | 60.24 (60.11, 60.37) | 63.22 (63.1, 63.34) | 67.39 (67.28, 67.5) | 69.23 (68.83, 69.63) | 75.0 (74.49, 75.51) | 49.35 (49.2, 49.5) | 85.56 (85.47, 85.64) |
| vicuna-7b-v1.5 | 100.0 (100.0, 100.0) | 0.0 (0.0, 0.0) | 97.59 (97.55, 97.63) | 97.7 (97.66, 97.74) | 96.74 (96.7, 96.78) | 82.05 (81.72, 82.38) | 100.0 (100.0, 100.0) | 64.94 (64.79, 65.08) | 64.44 (64.33, 64.56) |
| vicuna-7b-v1.5-w4-g128 | 100.0 (100.0, 100.0) | 0.0 (0.0, 0.0) | 96.39 (96.34, 96.44) | 97.7 (97.66, 97.74) | 98.91 (98.89, 98.94) | 94.87 (94.68, 95.06) | 100.0 (100.0, 100.0) | 90.91 (90.82, 91.0) | 83.33 (83.25, 83.42) |
| Llama-2-7b-chat-hf | 33.33 (28.53, 38.14) | 27.5 (27.13, 27.87) | 59.04 (58.9, 59.17) | 80.46 (80.36, 80.56) | 43.48 (43.36, 43.59) | 61.54 (61.12, 61.96) | 68.75 (68.2, 69.3) | 16.88 (16.77, 17.0) | 85.56 (85.47, 85.64) |
| Llama-2-7b-chat-hf-w4-g128 | 44.44 (39.38, 49.51) | 30.0 (29.62, 30.38) | 56.63 (56.49, 56.76) | 88.51 (88.43, 88.59) | 18.48 (18.39, 18.57) | 43.59 (43.16, 44.02) | 62.5 (61.93, 63.07) | 32.47 (32.33, 32.61) | 68.89 (68.78, 69.0) |
| vicuna-13b-v1.5 | 100.0 (100.0, 100.0) | 0.0 (0.0, 0.0) | 96.39 (96.34, 96.44) | 98.85 (98.82, 98.88) | 96.74 (96.7, 96.78) | 61.54 (61.12, 61.96) | 93.75 (93.46, 94.04) | 89.61 (89.52, 89.7) | 87.78 (87.7, 87.86) |
| vicuna-13b-v1.5-w4-g128 | 100.0 (100.0, 100.0) | 2.5 (2.37, 2.63) | 95.18 (95.12, 95.24) | 97.7 (97.66, 97.74) | 97.83 (97.79, 97.86) | 53.85 (53.42, 54.28) | 90.62 (90.28, 90.97) | 92.21 (92.13, 92.29) | 90.0 (89.93, 90.07) |
| WizardLM-13B-V1.2 | 55.56 (50.49, 60.62) | 52.5 (52.09, 52.91) | 83.13 (83.03, 83.23) | 80.46 (80.36, 80.56) | 83.7 (83.61, 83.78) | 17.95 (17.62, 18.28) | 18.75 (18.29, 19.21) | 67.53 (67.39, 67.67) | 76.67 (76.57, 76.77) |
| WizardLM-13B-V1.2-w4-g128 | 77.78 (73.54, 82.01) | 52.5 (52.09, 52.91) | 97.59 (97.55, 97.63) | 98.85 (98.82, 98.88) | 98.91 (98.89, 98.94) | 38.46 (38.04, 38.88) | 50.0 (49.41, 50.59) | 75.32 (75.2, 75.45) | 87.78 (87.7, 87.86) |
| Llama-2-13b-chat-hf | 33.33 (28.53, 38.14) | 10.0 (9.75, 10.25) | 36.14 (36.02, 36.27) | 33.33 (33.22, 33.45) | 42.39 (42.28, 42.5) | 51.28 (50.85, 51.71) | 21.88 (21.39, 22.36) | 42.86 (42.71, 43.01) | 55.56 (55.44, 55.67) |
| Llama-2-13b-chat-hf-w4-g128 | 33.33 (28.53, 38.14) | 17.5 (17.18, 17.82) | 74.7 (74.58, 74.82) | 63.22 (63.1, 63.34) | 55.43 (55.32, 55.55) | 76.92 (76.56, 77.29) | 53.12 (52.54, 53.71) | 38.96 (38.81, 39.11) | 73.33 (73.23, 73.44) |
| vicuna-33b-v1.3 | 100.0 (100.0, 100.0) | 25.0 (24.64, 25.36) | 97.59 (97.55, 97.63) | 98.85 (98.82, 98.88) | 83.7 (83.61, 83.78) | 89.74 (89.48, 90.01) | 84.38 (83.95, 84.8) | 80.52 (80.4, 80.64) | 98.89 (98.86, 98.91) |
| vicuna-33b-v1.3-w4-g128 | 100.0 (100.0, 100.0) | 45.0 (44.59, 45.41) | 97.59 (97.55, 97.63) | 98.85 (98.82, 98.88) | 88.04 (87.97, 88.12) | 97.44 (97.3, 97.57) | 84.38 (83.95, 84.8) | 85.71 (85.61, 85.82) | 97.78 (97.74, 97.81) |
| baseline | 100.0 (100.0, 100.0) | 100.0 (100.0, 100.0) | 100.0 (100.0, 100.0) | 100.0 (100.0, 100.0) | 100.0 (100.0, 100.0) | 97.44 (97.3, 97.57) | 87.5 (87.11, 87.89) | 88.31 (88.22, 88.41) | 96.67 (96.62, 96.71) |

**Table S11 (a). Model Accuracy_non-mentioned_ difference (compared to the baseline) on each SDoH question (Refined pipeline)**

| **Model** | **Q1** | **Q2** | **Q3** | **Q4** | **Q5** | **Q6** | **Q7** | **Q8** | **Q9** |
| --- | --- | --- | --- | --- | --- | --- | --- | --- | --- |
| openchat_3.5 | 0.0 (-0.0, 0.0) | -10.0 (-10.25, -9.75) | -4.82 (-4.88, -4.76) | -3.45 (-3.49, -3.4) | -2.17 (-2.21, -2.14) | 0.0 (-0.19, 0.19) | 0.0 (-0.54, 0.54) | 10.39 (10.29, 10.49) | 1.11 (1.06, 1.17) |
| openchat_3.5-w4-g128 | -11.11 (-14.31, -7.91) | -10.0 (-10.25, -9.75) | -4.82 (-4.88, -4.76) | -2.3 (-2.34, -2.26) | -2.17 (-2.21, -2.14) | -5.13 (-5.39, -4.86) | 0.0 (-0.54, 0.54) | 9.09 (8.98, 9.2) | 1.11 (1.06, 1.17) |
| zephyr-7b-beta | 0.0 (-0.0, 0.0) | -2.5 (-2.63, -2.37) | -7.23 (-7.3, -7.16) | -10.34 (-10.42, -10.27) | -15.22 (-15.3, -15.14) | -28.21 (-28.62, -27.79) | -3.12 (-3.69, -2.56) | -6.49 (-6.64, -6.34) | -2.22 (-2.29, -2.15) |
| zephyr-7b-beta-w4-g128 | 0.0 (-0.0, 0.0) | 0.0 (-0.0, 0.0) | -31.33 (-31.45, -31.2) | -34.48 (-34.6, -34.36) | -27.17 (-27.28, -27.07) | -12.82 (-13.16, -12.48) | -18.75 (-19.41, -18.09) | -14.29 (-14.45, -14.12) | -1.11 (-1.18, -1.05) |
| vicuna-7b-v1.5 | 0.0 (-0.0, 0.0) | -22.5 (-22.85, -22.15) | -1.2 (-1.23, -1.18) | -1.15 (-1.18, -1.12) | -2.17 (-2.21, -2.14) | 2.56 (2.43, 2.7) | 9.38 (8.94, 9.81) | -22.08 (-22.25, -21.91) | -32.22 (-32.34, -32.1) |
| vicuna-7b-v1.5-w4-g128 | 0.0 (-0.0, 0.0) | -37.5 (-37.9, -37.1) | -2.41 (-2.45, -2.37) | -1.15 (-1.18, -1.12) | 0.0 (-0.0, 0.0) | -2.56 (-2.8, -2.33) | 3.12 (2.61, 3.64) | 3.9 (3.77, 4.02) | -12.22 (-12.32, -12.13) |
| Llama-2-7b-chat-hf | -66.67 (-71.47, -61.86) | -17.5 (-17.82, -17.18) | -25.3 (-25.42, -25.18) | -11.49 (-11.57, -11.41) | -52.17 (-52.29, -52.06) | -28.21 (-28.62, -27.79) | -81.25 (-81.73, -80.77) | -58.44 (-58.61, -58.27) | -13.33 (-13.43, -13.24) |
| Llama-2-7b-chat-hf-w4-g128 | -55.56 (-60.62, -50.49) | -5.0 (-5.18, -4.82) | -40.96 (-41.1, -40.83) | -10.34 (-10.42, -10.27) | -79.35 (-79.44, -79.26) | -28.21 (-28.62, -27.79) | -75.0 (-75.54, -74.46) | -53.25 (-53.42, -53.08) | -27.78 (-27.89, -27.66) |
| vicuna-13b-v1.5 | 0.0 (-0.0, 0.0) | -10.0 (-10.25, -9.75) | -2.41 (-2.45, -2.37) | 0.0 (-0.0, 0.0) | -2.17 (-2.21, -2.14) | 2.56 (2.43, 2.7) | -65.62 (-66.24, -65.01) | 3.9 (3.77, 4.02) | -7.78 (-7.86, -7.69) |
| vicuna-13b-v1.5-w4-g128 | 0.0 (-0.0, 0.0) | -20.0 (-20.33, -19.67) | -3.61 (-3.66, -3.56) | -1.15 (-1.18, -1.12) | -1.09 (-1.11, -1.06) | 2.56 (2.43, 2.7) | -28.12 (-28.81, -27.44) | 6.49 (6.38, 6.61) | -5.56 (-5.63, -5.48) |
| WizardLM-13B-V1.2 | -44.44 (-49.51, -39.38) | -12.5 (-12.77, -12.23) | -16.87 (-16.97, -16.77) | -18.39 (-18.49, -18.29) | -15.22 (-15.3, -15.14) | -2.56 (-2.8, -2.33) | -3.12 (-3.69, -2.56) | -12.99 (-13.15, -12.83) | -18.89 (-19.0, -18.78) |
| WizardLM-13B-V1.2-w4-g128 | -22.22 (-26.46, -17.99) | -15.0 (-15.3, -14.7) | -2.41 (-2.45, -2.37) | -1.15 (-1.18, -1.12) | -1.09 (-1.11, -1.06) | -2.56 (-2.8, -2.33) | -21.88 (-22.55, -21.2) | -11.69 (-11.85, -11.53) | -8.89 (-8.98, -8.8) |
| Llama-2-13b-chat-hf | -66.67 (-71.47, -61.86) | -32.5 (-32.89, -32.11) | -48.19 (-48.33, -48.06) | -70.11 (-70.23, -70.0) | -50.0 (-50.11, -49.89) | -53.85 (-54.29, -53.4) | -59.38 (-60.02, -58.73) | -45.45 (-45.63, -45.28) | -24.44 (-24.56, -24.33) |
| Llama-2-13b-chat-hf-w4-g128 | -55.56 (-60.62, -50.49) | -35.0 (-35.4, -34.6) | -24.1 (-24.21, -23.98) | -36.78 (-36.9, -36.66) | -30.43 (-30.54, -30.33) | -46.15 (-46.6, -45.7) | -59.38 (-60.02, -58.73) | -36.36 (-36.54, -36.19) | -7.78 (-7.86, -7.69) |
| vicuna-33b-v1.3 | 0.0 (-0.0, 0.0) | -7.5 (-7.72, -7.28) | -2.41 (-2.45, -2.37) | 0.0 (-0.0, 0.0) | -16.3 (-16.39, -16.22) | 2.56 (2.43, 2.7) | -53.12 (-53.8, -52.45) | -5.19 (-5.34, -5.05) | 2.22 (2.17, 2.27) |
| vicuna-33b-v1.3-w4-g128 | 0.0 (-0.0, 0.0) | -7.5 (-7.72, -7.28) | -2.41 (-2.45, -2.37) | -1.15 (-1.18, -1.12) | -10.87 (-10.94, -10.8) | 2.56 (2.43, 2.7) | -59.38 (-60.02, -58.73) | -1.3 (-1.44, -1.16) | 2.22 (2.17, 2.27) |

**Table S11 (b). Model Accuracy_non-mentioned_ difference (compared to the baseline) on each SDoH question (Default pipeline)**

| **Model** | **Q1** | **Q2** | **Q3** | **Q4** | **Q5** | **Q6** | **Q7** | **Q8** | **Q9** |
| --- | --- | --- | --- | --- | --- | --- | --- | --- | --- |
| openchat_3.5 | 0.0 (-0.0, 0.0) | -37.5 (-37.9, -37.1) | -4.82 (-4.88, -4.76) | -3.45 (-3.49, -3.4) | -3.26 (-3.3, -3.22) | -46.15 (-46.6, -45.7) | 12.5 (12.11, 12.89) | 9.09 (8.98, 9.2) | 1.11 (1.06, 1.17) |
| openchat_3.5-w4-g128 | -11.11 (-14.31, -7.91) | -42.5 (-42.91, -42.09) | -4.82 (-4.88, -4.76) | -2.3 (-2.34, -2.26) | -3.26 (-3.3, -3.22) | -51.28 (-51.73, -50.83) | 12.5 (12.11, 12.89) | 7.79 (7.68, 7.9) | 1.11 (1.06, 1.17) |
| zephyr-7b-beta | -11.11 (-14.31, -7.91) | -12.5 (-12.77, -12.23) | -10.84 (-10.93, -10.76) | -13.79 (-13.88, -13.71) | -20.65 (-20.74, -20.56) | -10.26 (-10.57, -9.94) | 9.38 (8.94, 9.81) | -24.68 (-24.85, -24.5) | -4.44 (-4.52, -4.37) |
| zephyr-7b-beta-w4-g128 | 0.0 (-0.0, 0.0) | -12.5 (-12.77, -12.23) | -39.76 (-39.89, -39.63) | -36.78 (-36.9, -36.66) | -32.61 (-32.72, -32.5) | -28.21 (-28.62, -27.79) | -12.5 (-13.13, -11.87) | -38.96 (-39.14, -38.78) | -11.11 (-11.2, -11.02) |
| vicuna-7b-v1.5 | 0.0 (-0.0, 0.0) | -100.0 (-100.0, -100.0) | -2.41 (-2.45, -2.37) | -2.3 (-2.34, -2.26) | -3.26 (-3.3, -3.22) | -15.38 (-15.74, -15.03) | 12.5 (12.11, 12.89) | -23.38 (-23.55, -23.2) | -32.22 (-32.34, -32.1) |
| vicuna-7b-v1.5-w4-g128 | 0.0 (-0.0, 0.0) | -100.0 (-100.0, -100.0) | -3.61 (-3.66, -3.56) | -2.3 (-2.34, -2.26) | -1.09 (-1.11, -1.06) | -2.56 (-2.8, -2.33) | 12.5 (12.11, 12.89) | 2.6 (2.47, 2.73) | -13.33 (-13.43, -13.24) |
| Llama-2-7b-chat-hf | -66.67 (-71.47, -61.86) | -72.5 (-72.87, -72.13) | -40.96 (-41.1, -40.83) | -19.54 (-19.64, -19.44) | -56.52 (-56.64, -56.41) | -35.9 (-36.34, -35.46) | -18.75 (-19.41, -18.09) | -71.43 (-71.58, -71.28) | -11.11 (-11.2, -11.02) |
| Llama-2-7b-chat-hf-w4-g128 | -55.56 (-60.62, -50.49) | -70.0 (-70.38, -69.62) | -43.37 (-43.51, -43.24) | -11.49 (-11.57, -11.41) | -81.52 (-81.61, -81.43) | -53.85 (-54.29, -53.4) | -25.0 (-25.68, -24.32) | -55.84 (-56.01, -55.67) | -27.78 (-27.89, -27.66) |
| vicuna-13b-v1.5 | 0.0 (-0.0, 0.0) | -100.0 (-100.0, -100.0) | -3.61 (-3.66, -3.56) | -1.15 (-1.18, -1.12) | -3.26 (-3.3, -3.22) | -35.9 (-36.34, -35.46) | 6.25 (5.77, 6.73) | 1.3 (1.17, 1.43) | -8.89 (-8.98, -8.8) |
| vicuna-13b-v1.5-w4-g128 | 0.0 (-0.0, 0.0) | -97.5 (-97.63, -97.37) | -4.82 (-4.88, -4.76) | -2.3 (-2.34, -2.26) | -2.17 (-2.21, -2.14) | -43.59 (-44.04, -43.14) | 3.12 (2.61, 3.64) | 3.9 (3.77, 4.02) | -6.67 (-6.75, -6.58) |
| WizardLM-13B-V1.2 | -44.44 (-49.51, -39.38) | -47.5 (-47.91, -47.09) | -16.87 (-16.97, -16.77) | -19.54 (-19.64, -19.44) | -16.3 (-16.39, -16.22) | -79.49 (-79.84, -79.13) | -68.75 (-69.34, -68.16) | -20.78 (-20.95, -20.61) | -20.0 (-20.11, -19.89) |
| WizardLM-13B-V1.2-w4-g128 | -22.22 (-26.46, -17.99) | -47.5 (-47.91, -47.09) | -2.41 (-2.45, -2.37) | -1.15 (-1.18, -1.12) | -1.09 (-1.11, -1.06) | -58.97 (-59.41, -58.53) | -37.5 (-38.2, -36.8) | -12.99 (-13.15, -12.83) | -8.89 (-8.98, -8.8) |
| Llama-2-13b-chat-hf | -66.67 (-71.47, -61.86) | -90.0 (-90.25, -89.75) | -63.86 (-63.98, -63.73) | -66.67 (-66.78, -66.55) | -57.61 (-57.72, -57.5) | -46.15 (-46.6, -45.7) | -65.62 (-66.24, -65.01) | -45.45 (-45.63, -45.28) | -41.11 (-41.24, -40.99) |
| Llama-2-13b-chat-hf-w4-g128 | -66.67 (-71.47, -61.86) | -82.5 (-82.82, -82.18) | -25.3 (-25.42, -25.18) | -36.78 (-36.9, -36.66) | -44.57 (-44.68, -44.45) | -20.51 (-20.9, -20.13) | -34.38 (-35.07, -33.68) | -49.35 (-49.53, -49.18) | -23.33 (-23.45, -23.22) |
| vicuna-33b-v1.3 | 0.0 (-0.0, 0.0) | -75.0 (-75.36, -74.64) | -2.41 (-2.45, -2.37) | -1.15 (-1.18, -1.12) | -16.3 (-16.39, -16.22) | -7.69 (-7.98, -7.4) | -3.12 (-3.69, -2.56) | -7.79 (-7.94, -7.64) | 2.22 (2.17, 2.27) |
| vicuna-33b-v1.3-w4-g128 | 0.0 (-0.0, 0.0) | -55.0 (-55.41, -54.59) | -2.41 (-2.45, -2.37) | -1.15 (-1.18, -1.12) | -11.96 (-12.03, -11.88) | 0.0 (-0.19, 0.19) | -3.12 (-3.69, -2.56) | -2.6 (-2.74, -2.46) | 1.11 (1.06, 1.17) |

**Table S12. Examples of invalid responses of each LLM on the refined pipeline (if applicable)**

| **Model** | **SDoH Question** | **Human Label** | **Model Response** | **Comments** |
| --- | --- | --- | --- | --- |
| openchat_3.5-w4-g128 | Q8. What is the patient’s highest education level? | College | The patient's highest education level is "Associates". This is not one of the options you provided, but it is a level of education between high school and a bachelor's degree. It is typically obtained after completing a two-year program at a community college or technical school. | The model correctly found the key information and gave strong reasoning. Since we did not specify “Associates” can be considered as “College” in the prompt. It is understandable that the model did not get this. |
| zephyr-7b-beta | Q1. What is the patient’s marital status? | Married | Certainly! Here are some common symptoms of alcohol abuse:  1. Drinking more or for longer periods than intended  2. Persistent desire or unsuccessful efforts to cut down or control alcohol use  3. Spending a lot of time obtaining, using, or recovering from the effects of alcohol  4. Cravings or strong urges to drink  5. Failing to fulfill obligations at work, school, or home due to alcohol use  6. Continuing to use alcohol despite physical or psychological problems caused or worsened by alcohol use  7. Giving up or reducing important social, occupational, or recreational activities because of alcohol use  8. Using alcohol in dangerous situations, such as while driving or operating machinery  9. Continuing to use alcohol despite knowledge of having a physical or psychological problem that is likely to have been caused or worsened by alcohol use  10. Tolerance, meaning that you need more alcohol to feel the same effect as before or that your usual amount of alcohol has less effect than before.  I hope this helps! Let me know if you have any other questions. | The model is answering the symptoms of alcohol abuse, which is totally irrelevant to the question. |
| zephyr-7b-beta-w4-g128 | Q7. What is the patient’s employment status? | Retired | Based on the provided employment information, it seems that the patient's employment status is "Jobless". While the patient previously held the position of "administrative assistant at a school", there is no clear indication provided as to whether this employment is current or not. Without further information, it is unclear whether the patient is currently employed, retired, or jobless. | “Retired” and “Jobless” are usually difficult to distinguish and the description about the patient’s employment status is usually ambiguous in the text. It is understandable that the model prefers “Jobless” over “Retired”. |
| vicuna-7b-v1.5 | Q9. Does the patient exercise? | Not mentioned | Based on the information provided, it cannot be determined whether the patient exercises or not. The information provided does not mention the patient's exercise habits or physical activity level. | The model gets this one correctly, but the post-processing failed to extract the answer. |
| vicuna-7b-v1.5-w4-g128 | Q7. What is the patient’s employment status? | Employed | Based on the given text, the patient's employment status is "Not retired as of yet golfs every day" which means the patient is not retired and is still working. Therefore, the answer to the question "What is the patient’s employment status?" is "Employed". | The model gets this one correctly. However, since both “retired” and “employed” appear in the response, the post-processing procedure fails to map this response to one of the choices. |
| Llama-2-7b-chat-hf | Q6. Does the patient live alone? | Not mentioned | You're welcome! It was my pleasure to assist you. If you have any other questions or concerns in the future, don't hesitate to reach out. Have a great day! | The model did not answer the question but replied with a standard ending sentence. |
| Llama-2-7b-chat-hf-w4-g128 | Q3. Does the patient currently use tobacco? | Not mentioned | Thank you for asking. I'm just an AI, I don't have access to real-time information about the patient's current tobacco use. However, I can tell you that it is important to be honest and accurate when reporting a patient's medical history, including their tobacco use. (p) N .  If the patient does not currently use tobacco, it is important to note that they may have used it in the past or be at risk of starting in the future. As a healthcare professional, it is crucial to assess the patient's overall health and lifestyle habits to provide appropriate care and recommendations. (p) N .  If you have any further questions or concerns, please feel free to ask. I'm here to help. (p) N | The model is not answering the question. The information in the response is not useful. |
| vicuna-13b-v1.5 | Q8. What is the patient’s highest education level? | High school | 12 is not a valid answer for the patient's highest education level. Please provide a valid answer from the given options. | The model precisely found that the patient has 12 years of education, but it fails to convert this to one of the choices. |
| vicuna-13b-v1.5-w4-g128 | Q8. What is the patient’s highest education level? | Not mentioned | 2 Years Education is not sufficient information to determine the patient's highest education level. | The “2” that the model found is not related to the years of education, but related to the number of children that the patient has. |
| WizardLM-13B-V1.2 | Q9. Does the patient exercise? | Not mentioned | 1. Does the patient exercise?  - Yes  Rationale: The text does not provide explicit information about the patient's current exercise habits. However, it does mention that the patient is retired and used to work as a medical assistant, which suggests that they may have had a more active lifestyle in the past. Therefore, it is reasonable to assume that the patient may still exercise, but without specific information, the answer is "Yes.".  - Yes | The model is using unfounded assumptions instead of the evidence in the text to infer an answer. |
| WizardLM-13B-V1.2-w4-g128 | Q2. How many children does the patient have? | 5 or more | 5  The patient has 5 children. | The model gets the answer correctly, but since the response does not contain “5 or more”. No answer is matched in the post-processing procedure. |
| Llama-2-13b-chat-hf | Q4. Does the patient currently consume alcohol? | Not mentioned | You're welcome! It was my pleasure to assist you. If you have any more questions or need further clarification on any other information, please don't hesitate to ask. Have a great day! | The model did not answer the question but replied with a standard ending sentence. |
| Llama-2-13b-chat-hf-w4-g128 | Q5. Does the patient currently use illicit drugs? | Not mentioned | You're welcome! It's my pleasure to assist you. If you have any other questions, feel free to ask. Have a great day! | The model did not answer the question but replied with a standard ending sentence. |
| vicuna-33b-v1.3 | Q7. What is the patient’s employment status? | Employed | 1. What is the patient's employment status?  a. Employed  Explanation: The text mentions that the patient is not retired as of yet and golfs every day, which implies that the patient has leisure time but does not explicitly mention a job. However, since the patient is not retired, it is reasonable to assume that they have an employed status. | The model gets the correct answer, but since both “employed” and “retired” are in the model response, the post-processing procedure cannot map the response to a single choice. |
